# Preconception indicators and associations with health outcomes reported in UK routine primary care data: a systematic review

**DOI:** 10.1101/2024.02.05.24302342

**Authors:** Danielle Schoenaker, Elizabeth M Lovegrove, Emma H Cassinelli, Jennifer Hall, Majel McGranahan, Laura McGowan, Helen Carr, Nisreen A Alwan, Judith Stephenson, Keith M Godfrey

## Abstract

**Background:** Routine primary care data may be a valuable resource for preconception health research and informing provision of preconception care.

**Aim:** To review how primary care data could provide information on the prevalence of preconception indicators and examine associations with maternal and offspring health outcomes.

**Design and Setting:** Systematic review of observational studies using UK routine primary care data.

**Method:** Literature searches were conducted in five databases (March 2023) to identify observational studies that used national primary care data from individuals aged 15-49 years. Preconception indicators were defined as medical, behavioural and social factors that may impact future pregnancies. Health outcomes included those that may occur during and after pregnancy. Screening, data extraction and quality assessment were conducted by two reviewers.

**Results:** From 5,259 records screened, 42 articles were included. The prevalence of 30 preconception indicators was described for female patients, ranging from 0.01% for sickle cell disease to >20% for each of advanced maternal age, previous caesarean section (among those with a recorded pregnancy), overweight, obesity, smoking, depression and anxiety (irrespective of pregnancy). Few studies reported indicators for male patients (n=3) or associations with outcomes (n=5). Most studies had low risk of bias, but missing data may limit generalisability.

**Conclusion:** Findings demonstrate that routinely collected UK primary care data can be used to identify patients’ preconception care needs. Linking primary care data with health outcomes collected in other datasets is underutilised but could help quantify how optimising preconception health and care can reduce adverse outcomes for mothers and children.

**How this fits in:**

- Provision of preconception care is not currently embedded into routine clinical practice but may be informed by routinely collected primary care data.
- This systematic review demonstrates that UK primary care data can provide information on the prevalence of a range of medical, behavioural and social factors among female patients of reproductive age, while limited research has examined male preconception health or associations with maternal and offspring health outcomes.
- Routinely recorded electronic patient record data can be used by primary healthcare professionals to search for preconception risk factors and thereby support individualised preconception care, while aggregate data can be used by public health agencies to promote population-level preconception health.
- Further data quality improvements and linkage of routine health datasets are needed to support the provision of preconception care and future research on its benefits for maternal and offspring health outcomes.

## Introduction

Preconception care is the provision of biomedical, behavioural and social interventions to people of reproductive age (15-49 years) before conception may occur with the aim of improving short-and longer-term parental and child health outcomes.^1^ Primary care teams have a key role in providing preconception care as identified by patients and healthcare professionals.^2, 3^ Preconception care delivered in primary care improves knowledge and preconception health behaviours in female patients, but there is currently less evidence about male patients or the impact on pregnancy and longer-term health outcomes.^4, 5^ In line with the National Institute for Health and Care Excellence (NICE) Clinical Knowledge Summary on preconception advice and management, primary care teams are encouraged to consider discussions about preconception health when appropriate, and to assess, manage and potentially optimise a range of physical and mental health conditions, health behaviours, and social needs prior to potential pregnancy.^6^ However, routine provision of preconception care is not currently widespread in UK clinical practice.^7^

To build the case for implementation of strategies and guidelines that optimise the population’s preconception health, the UK Preconception Partnership proposed an annual report card to describe and monitor preconception health.^8^ Our scoping review to inform national surveillance identified 65 preconception indicators (medical, behavioural and social risk factors that may impact potential future pregnancies among individuals of reproductive age) that are recorded in existing UK routine health data.^9^ A first report card was produced based on 23 indicators recorded in the national Maternity Service Data Set (MSDS), demonstrating that nine in 10 women in England enter pregnancy with at least one potentially modifiable risk factor for adverse pregnancy and birth outcomes.^10, 11^ Similarly, an analysis of primary care data from the Royal College of General Practitioners Research and Surveillance Centre found that 91% of women of reproductive age have a behavioural or medical risk factor for adverse pregnancy outcomes.^12^ These studies have to date focussed on preconception health of women (not men), and have not examined trends and trajectories in medical, behavioural and social indicators during the years leading up to pregnancy. Doing so would improve our ability to identify the population’s preconception care needs throughout their reproductive years. Routinely collected primary care data is potentially a unique resource to describe and monitor preconception health, and to examine the impact of (changes in) preconception indicators on improving outcomes such as gestational diabetes and preterm birth.

To inform future research and surveillance, and develop policy and clinical practice recommendations, we aimed to systematically review the literature to explore how UK routine primary care data could provide information on the prevalence of preconception indicators and examine associations with maternal and offspring health outcomes.

## Methods

### Search strategy and selection criteria

The protocol for this review was registered with PROSPERO,^13^ and the Preferred Reporting Items for Systematic reviews and Meta-Analyses (PRISMA 2020) guideline used to ensure transparent reporting.^14^ A search strategy was developed, and searches conducted on 27 March 2023 (from inception date) in five databases: MEDLINE (Ovid), EMBASE (Ovid), Scopus, CINAHL, Web of Science (Supplementary Table 1). Supplementary searches using ‘preconception’ and ‘prepregnancy’ terms were conducted using databases from the British Journal of General Practice, and UK primary care datasets.^13^ Reference lists of included articles were screened for additional studies.

Articles were selected if they included findings from an observational study among individuals of reproductive age (15-49 years), used national patient-level routine primary care data collected in England, Wales, Scotland and/or Northern Ireland, and reported on the prevalence of at least one preconception indicator identified from our previous scoping review (Table 1).^9^ Articles not including new/original peer-reviewed results, and conference abstracts, were excluded.

**Table 1.** PICOS statement.

|  |  |
| --- | --- |
| Population | Individuals of reproductive age who may or may not be(come) pregnant/conceive a pregnancy (any gender, aged 15-49 years). |
| Intervention/<br>exposure | Preconception indicators as identified in Schoenaker et al.<sup>9</sup><br>Preconception indicators are defined as medical, behavioural and social risk factors or exposures as well as wider determinants of health that may impact potential future pregnancies among all individuals of reproductive age. Studies do not have to identify relevant factors or exposures as 'preconception indicators'. |
| Comparator/<br>control | Not applicable. |
| Outcome | Maternal health outcomes: any outcome that may occur during pregnancy (e.g. gestational diabetes), delivery (e.g. caesarean section), postpartum (e.g. mortality), or beyond (no age limit) (e.g. type 2 diabetes).<br>Offspring health and developmental outcomes (including social/educational outcomes): any outcome that may occur during pregnancy (e.g. stillbirth), delivery (e.g. preterm birth), infancy (e.g. neonatal intensive care unit admission), or beyond (no age limit) (e.g. learning difficulty). |
| Study design | Observational studies (including cohort, cross-sectional and case-control studies). |

### Selection process

Search results were collated in EndNote and duplicates removed, before uploading to Covidence software. Titles and abstracts, followed by full text articles, were screened independently by two reviewers for inclusion. Disagreements or uncertainties were resolved through discussion.

### Data extraction and synthesis

A standardised data extraction form was developed and piloted. Data were extracted by one reviewer and checked by a second reviewer. Disagreements were resolved between the two reviewers. All extracted data on study characteristics (grouped by primary care database), prevalence of preconception indicators, and measures of association between preconception indicators and outcomes (grouped by preconception indicator), were presented in tables. Meta-analysis was not conducted due to heterogeneity in preconception indicator definitions and inclusion and exclusion criteria of study populations.

### Risk of bias assessment

Risk of bias was assessed for study findings on the prevalence of preconception indicators using the 10-item scale developed by Hoy et al rating internal and external validity.^15^ The Newcastle-Ottawa Scale (NOS) was used to rate risk of bias of study findings on associations between preconception indicators and health outcomes based on seven items related to selection, comparability, and exposure/outcome.^16^ Risk of bias was assessed by one reviewer, and checked by a second reviewer. Disagreements were resolved between the two reviewers. Studies were classified as low, moderate or high risk of bias (findings on prevalence),^15^ and good, fair or poor data quality (findings on associations)^16^ (scoring guides in Supplementary Tables 2, 3A-3B).

## Results

From 9,401 identified records, 4,142 duplicates were removed and after title and abstract screening (n=5,259), 117 full-text articles were evaluated for eligibility (Figure 1). 42 articles were included, reporting findings from 11 primary care databases such as the Clinical Practice Research Datalink (CPRD) and The Health Improvement Network (THIN).

**Figure 1.**
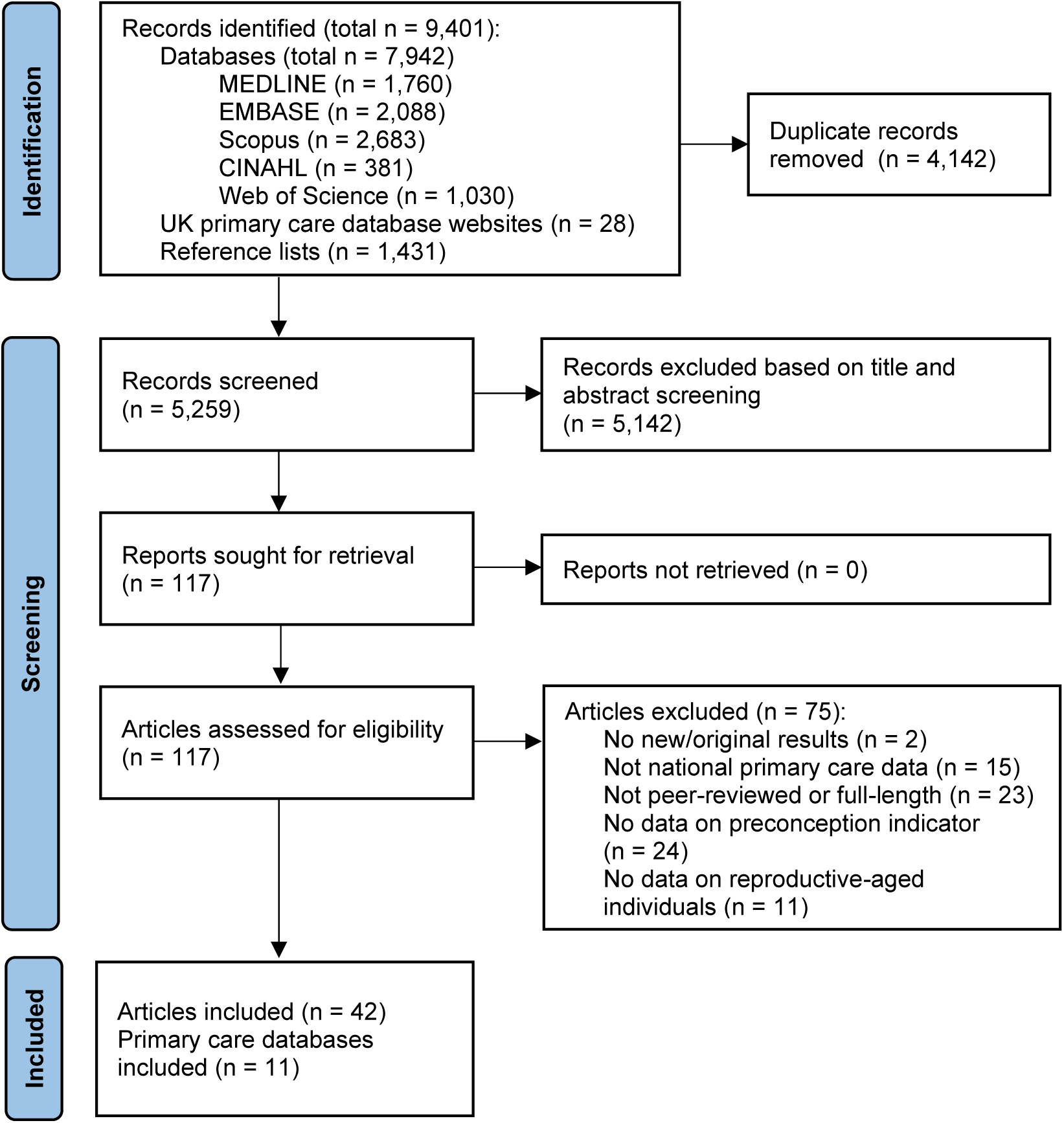
PRISMA flow diagram for the identification and selection of studies included in the review.

Most articles reported findings from primary care databases that included patients from three (n=1) or all four UK nations (n=30), or from England (n=6), Scotland (n=3) or Northern Ireland only (n=2) (Table 2, Supplementary Table 4). In 11 studies, a primary care dataset was linked with at least one other dataset, such as Hospital Episodes Statistics (HES), Office for National Statistics (ONS) mortality register, community prescribing data, or the Avon Longitudinal Study of Parents and Children. All studies included data on female patients; three studies also reported preconception indicators for male patients.

**Table 2.** Characteristics of included studies reporting on the prevalence of preconception indicators in the overall population of people of reproductive age.

| First author, year (reference) | Dataset | Country | Study design | Data collection period | Total or maximum sample size | Population characteristics: sex, age | Preconception indicators reported | Maternal and offspring outcomes reported |
| --- | --- | --- | --- | --- | --- | --- | --- | --- |
| ***Clinical Practice Research Datalink (CPRD)*** |  |  |  |  |  |  |  |  |
| Lee, 2022 <sup>(27)</sup> | CPRD GOLD | UK | Cross-sectional study | 2018 | 37,641 | Female, 15-49 years | Maternal age, ethnicity, deprivation, weight, smoking, depression, anxiety, severe mental health condition, asthma, PCOS, infertility, thyroid disease, eating disorder, endometriosis, hypertension, thromboembolism, sickle-cell disease, epilepsy, diabetes | None |
| Hope, 2022 <sup>(35)</sup> | CPRD GOLD | UK | Cohort study | 1990-2017 | 2,680,149 | Female, 14-45 years | Ethnicity, mental health condition | None |
| Subramanian, 2022 <sup>(19)</sup> | CPRD GOLD linked with HES | England | Cohort and case-control study | 1997-2020 | 299,866 | Female, 15-49 years | Ethnicity, PCOS | Preterm delivery, mode of delivery, high or low birthweight, stillbirth, small and large for gestational age |
| Syed, 2022 <sup>(36)</sup> | CPRD GOLD linked with HES and Office for National Statistics (ONS) mortality register | England | Cohort study | 2004- 2018 | 211,393 | Female, 16-55 years | Deprivation, ethnicity, maternal age | None |
| den Heijer, 2019 <sup>(25)</sup> | CPRD GOLD linked with IMD | England | Cohort | 2000-2013 | 857,324 | Female, 12-25 years | Smoking, deprivation, folic acid supplementation, sexually transmitted disease, PCOS, endometriosis, thyroid disease | Ectopic pregnancy |
| ***General Practice Research Database (GPRD)*** |  |  |  |  |  |  |  |  |
| Briggs, 2013 <sup>(37)</sup> | N/A | UK | Repeated cross-sectional study | 2004-2010 | 1,103,669 | Female, 15-49 years | Contraception | None |
| Rowlands, 2000 <sup>(38)</sup> | N/A | UK | Cohort study | 1994-1997 | 95,007 | Female, 14-29 years | Contraception | None |
| ***The Health Improvement Network (THIN)*** |  |  |  |  |  |  |  |  |
| Smith, 2020 <sup>(39)</sup> | N/A | UK | Cohort study | 2006-2016 | 241,662 | Female, 15-49 years | Maternal age, deprivation, previous caesarean delivery, smoking | None |
| Cea Soriano, 2018 <sup>(40)</sup> | N/A | UK | Cohort study and case-control study | 1995-2012 | 251,581 | Female, 15-45 years | Diabetes mellitus | None |
| Coton, 2016 <sup>(30)</sup> | N/A | UK | Cohort study | 1995-2012 | 301,794 | Female, 16 and over | Diabetes mellitus | None |
| Ban, 2015 (2) <sup>(41)</sup> | N/A | UK | Cohort study | 1990-2010 | 2,141,503 | Female, 15-44 years | Inflammatory bowel disease | None |
| Cea Soriano, 2014 <sup>(28)</sup> | N/A | UK | Cohort study | 2004-2010 | N/R | Female, 18-44 years | Contraception | None |
| Cea Soriano, 2013 <sup>(42)</sup> | N/A | UK | Cohort study | 2008 | 574,185 | Female, 12-49 years | Contraception | None |
| Dhalwani, 2013 <sup>(43)</sup> | N/A | UK | Cohort study | 1990-2010 | 1,776,746 | Female, 15-49 years | Fertility problems | None |
| Ban, 2012 <sup>(44)</sup> | N/A | UK | Cohort study | 1994-2009 | 116,457 | Female, 15-45 years | Depression, anxiety, serious mental illness, deprivation | None |
| Dave, 2010 <sup>(17)</sup> | Use of family identification number to link mothers, fathers and children living in the same household | UK | Cohort study | 1993-2007 | 86,957 | Female and male, 15-≥35 years | Depression | None |
| ***Royal College of General Practitioners (RCGP) Research and Surveillance Centre (RSC) network*** |  |  |  |  |  |  |  |  |
| Gaudio, 2022 <sup>(18)</sup> | N/A | England | Repeated cross-sectional study | 2004-2018 | 729,662 | Female and male, 12-46 years | Valproate prescription | None |
| Gaudio, 2021 <sup>(29)</sup> | N/A | England | Repeated cross-sectional study | 2004-2017 | 465,898 | Female, 16-45 years | Diabetes mellitus | None |
| **Enhanced Prescribing Database (EPD)** |  |  |  |  |  |  |  |  |
| Given, 2020 <sup>(45)</sup> | EPD linked with GP Patient Registrations Index | Northern Ireland | Cohort study | 2010-2016 | 560,074 | Female, 12-49 years | Contraception | None |
| Wemakor, 2014 <sup>(46)</sup> | EPD linked with the 2010 Northern Ireland Multiple Deprivation Measure (NIMDM) | Northern Ireland | Cross-sectional study | 2009 | 268,917 | Female, 15-45 years | Deprivation, anti-depressant use | None |
| **Other datasets** |  |  |  |  |  |  |  |  |
| Lee, 2022 <sup>(27)</sup> | Secure Anonymised Information Linkage (SAIL) Databank | Wales | Cross-sectional study | 2018 | 27,782 | Female, 15-49 years | Maternal age, ethnicity, deprivation, weight, smoking, mental health problem, severe mental health condition, asthma, PCOS, infertility, thyroid disease, eating disorder, endometriosis, hypertension, thromboembolism, sickle-cell disease, epilepsy, diabetes | None |
| Pasvol, 2022 <sup>(47)</sup> | IQVIA Medical Research Data (IMRD) database | UK | Repeated cross-sectional study | 2000-2018 | 2,705,638 | Female, 15-49 years | Contraception | None |

ALSPAC, Avon Longitudinal Study of Parents and Children; CPRD, Clinical Practice Research Datalink; EPD, Enhanced Prescribing Database; HES, Hospital Episodes Statistics; IMD, index of multiple deprivation; PCOS, polycystic ovary syndrome; SD, standard deviation.

### Prevalence of preconception indicators

Articles reported findings on 30 preconception indicators across seven of the 12 domains identified in our scoping review.^9^ Most studies included people of reproductive age irrespective of past/future pregnancy (n=26), while other studies included women with a pregnancy or birth recorded during the study period (n=15) or women with a recorded pregnancy and their partners (n=1) (Supplementary Table 5).

To obtain population-level estimates of preconception indicators, prevalence data were extracted only if reported (or could be calculated) for the overall study population of females or males of reproductive age (i.e. not if reported only in sub-populations such as patients with a specific condition or characteristic) (Table 3). Data on overall prevalence were available for 21 of the 42 studies, with the other 21 studies reporting prevalence estimates only in sub-populations. Additional preconception indicators reported in sub-populations included housing, domestic abuse, routine GP check-up in the past year, paternal age, previous pregnancy loss, history of assisted reproduction, alcohol consumption, substance misuse, cervical screening, and cardiovascular disease (Supplementary Table 4).

**Table 3.**
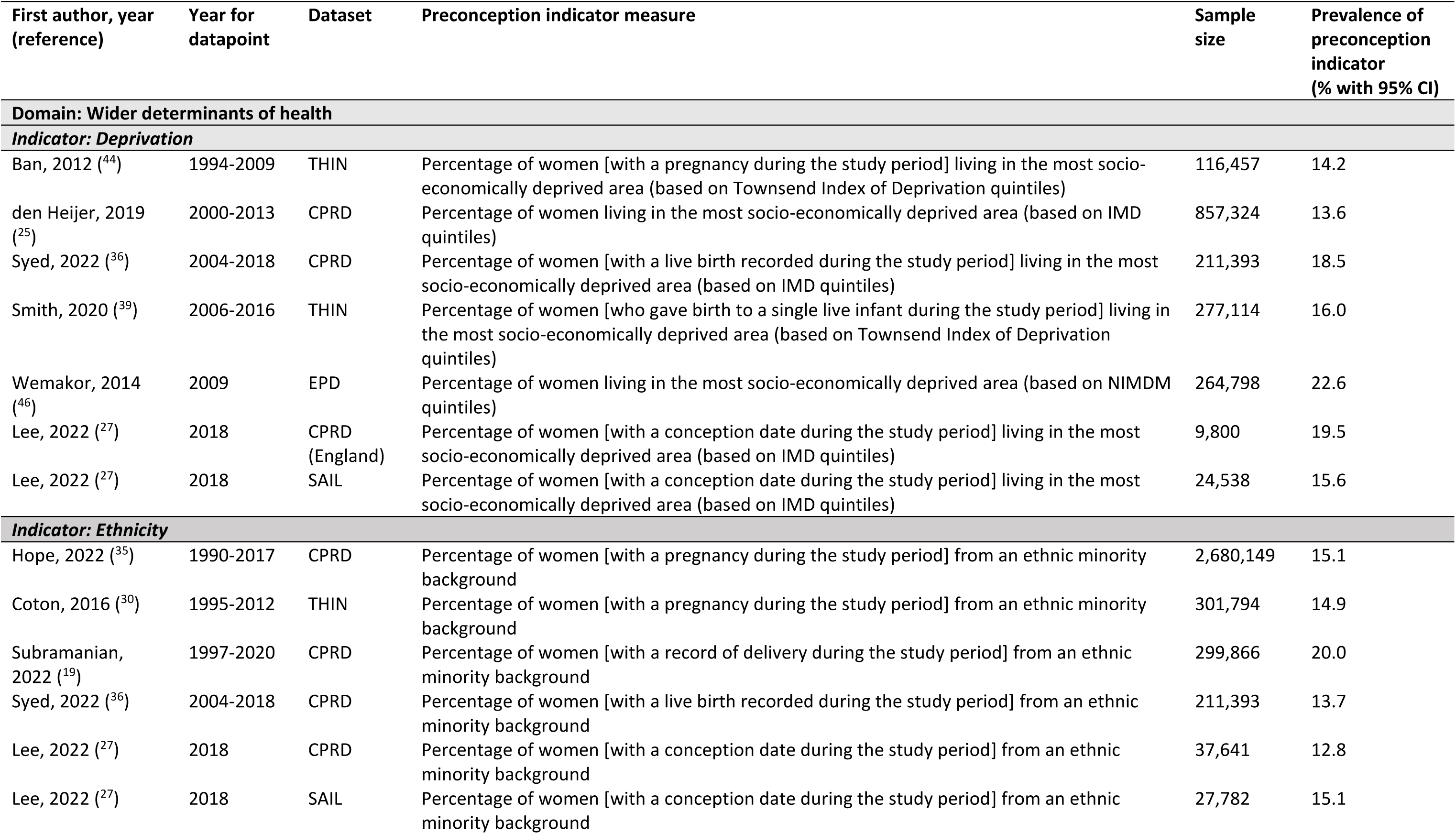
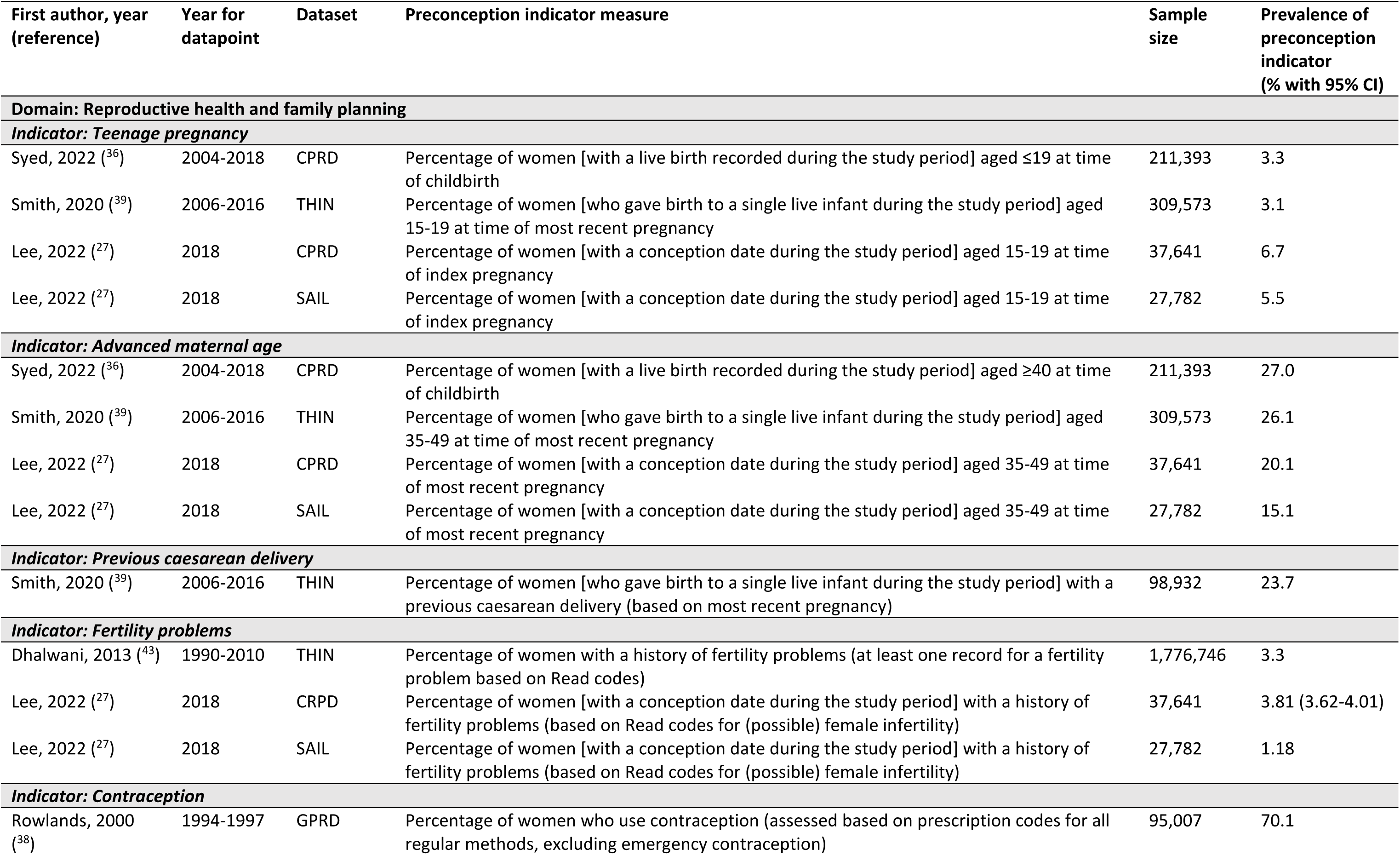
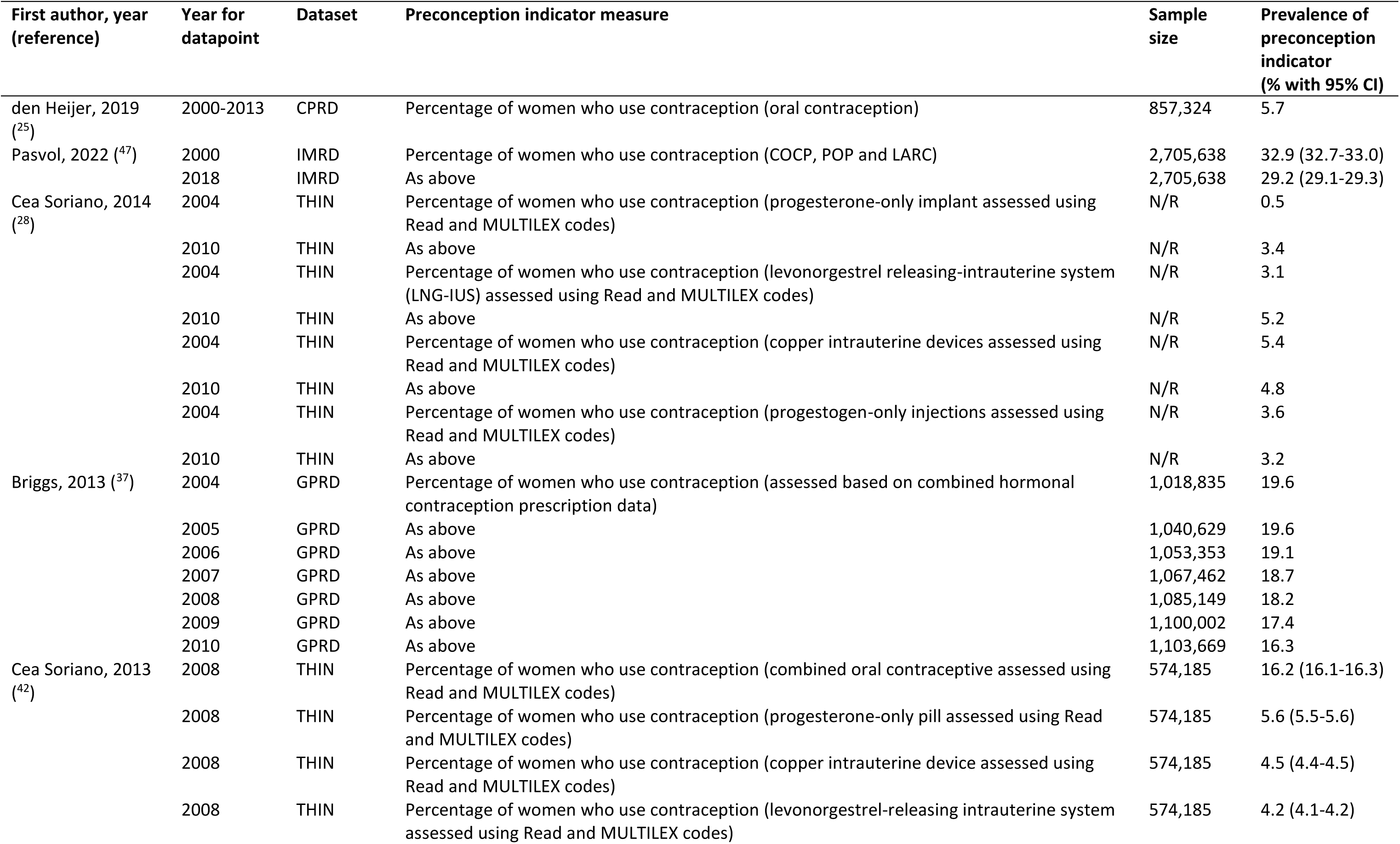
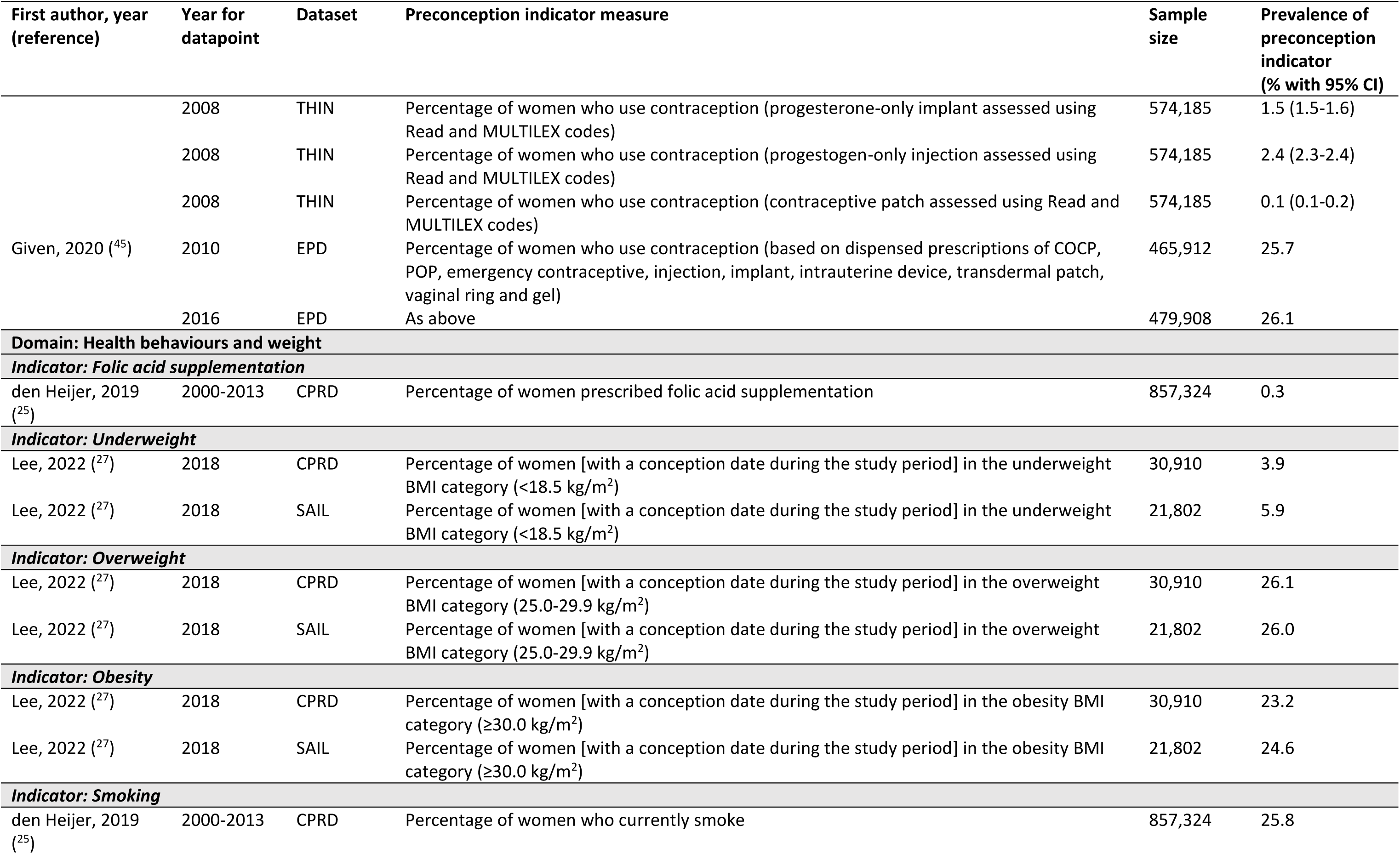
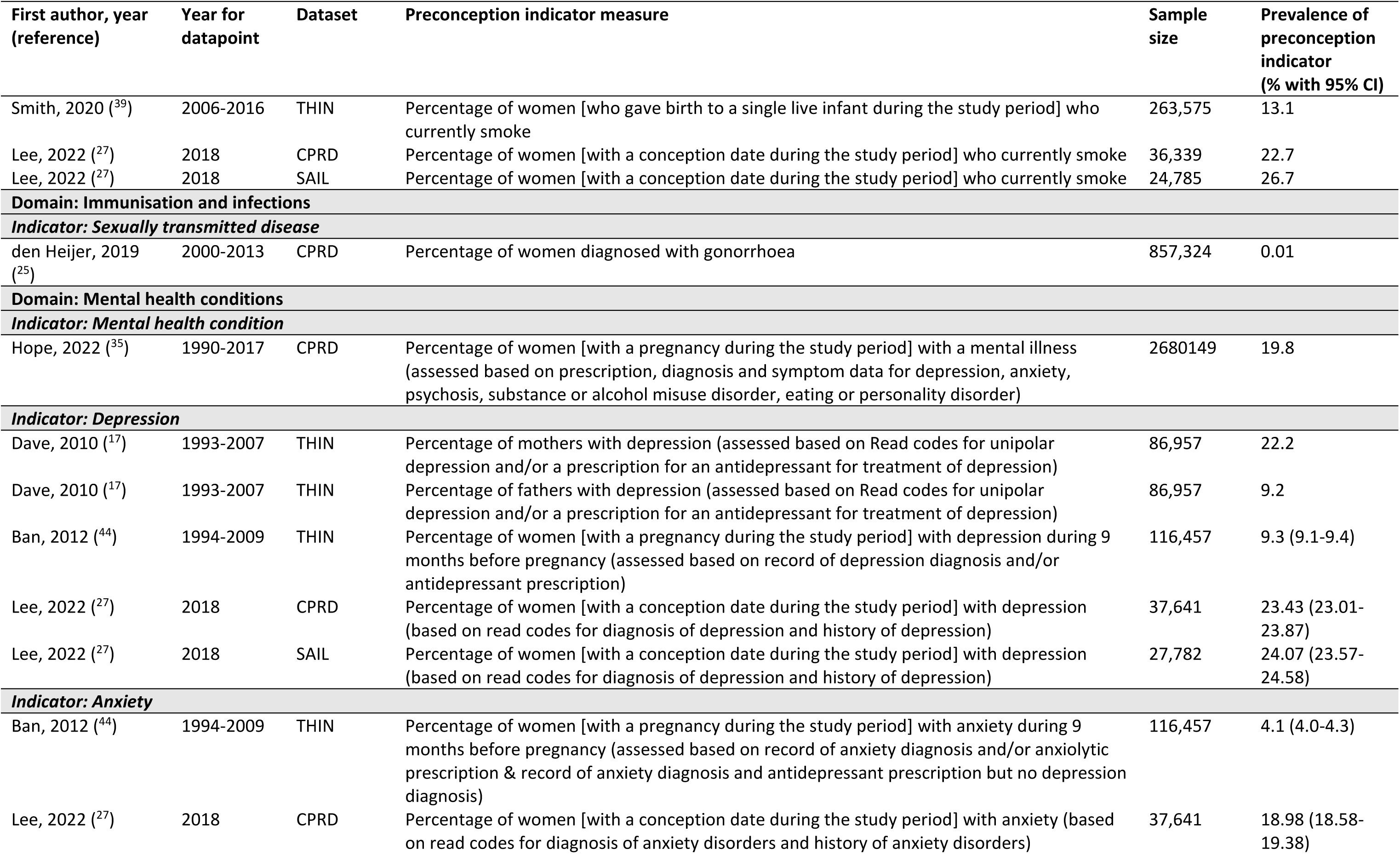
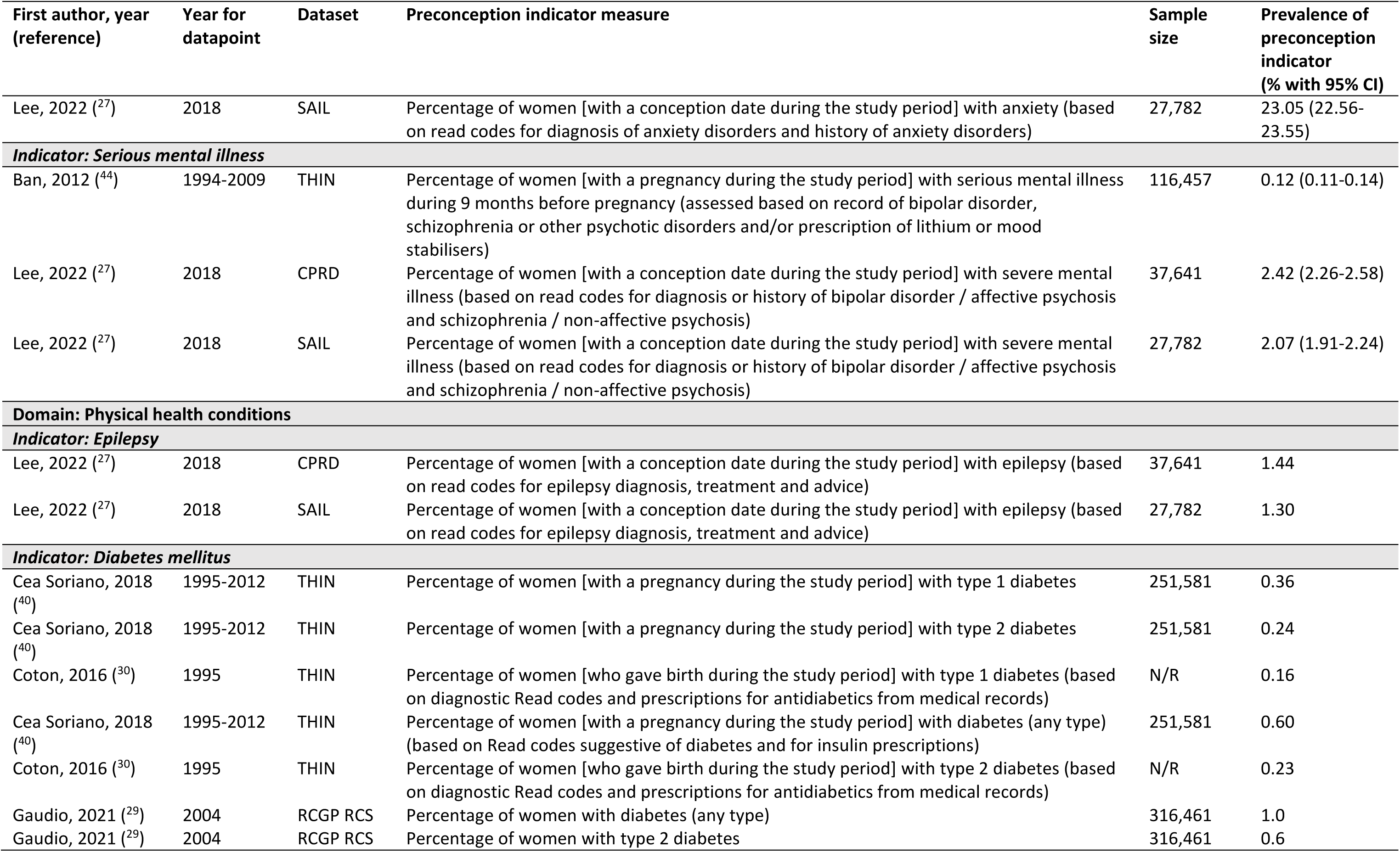
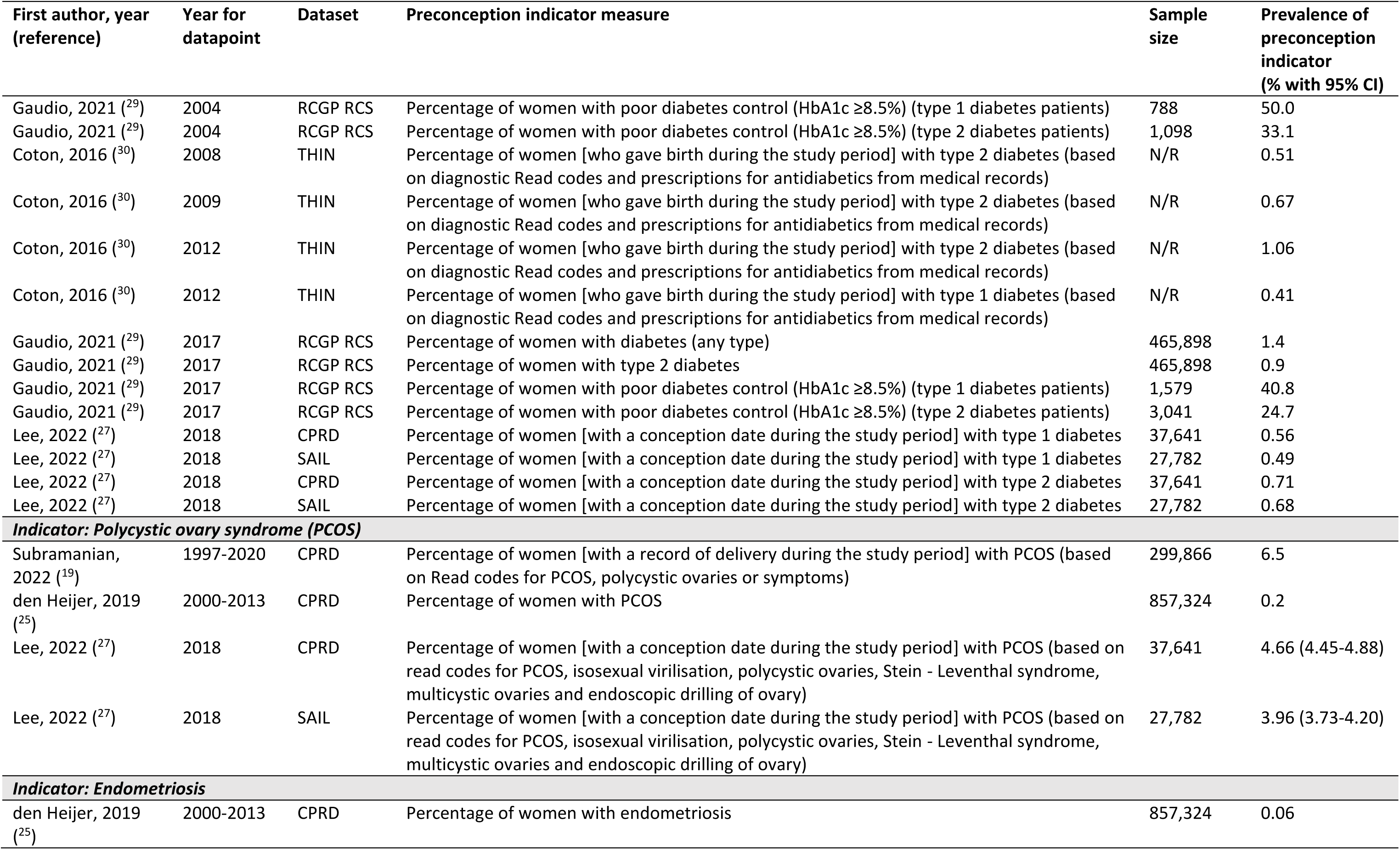
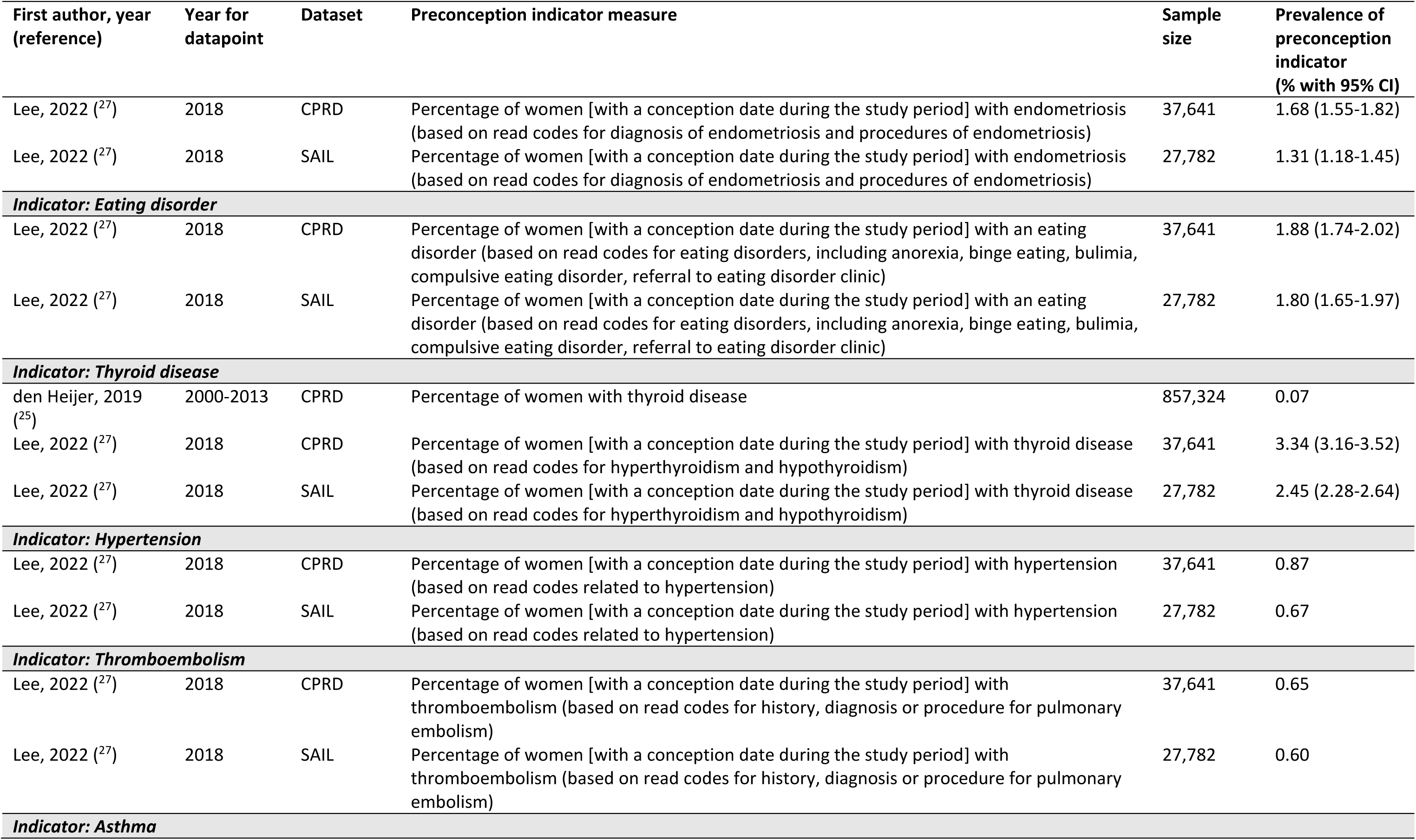
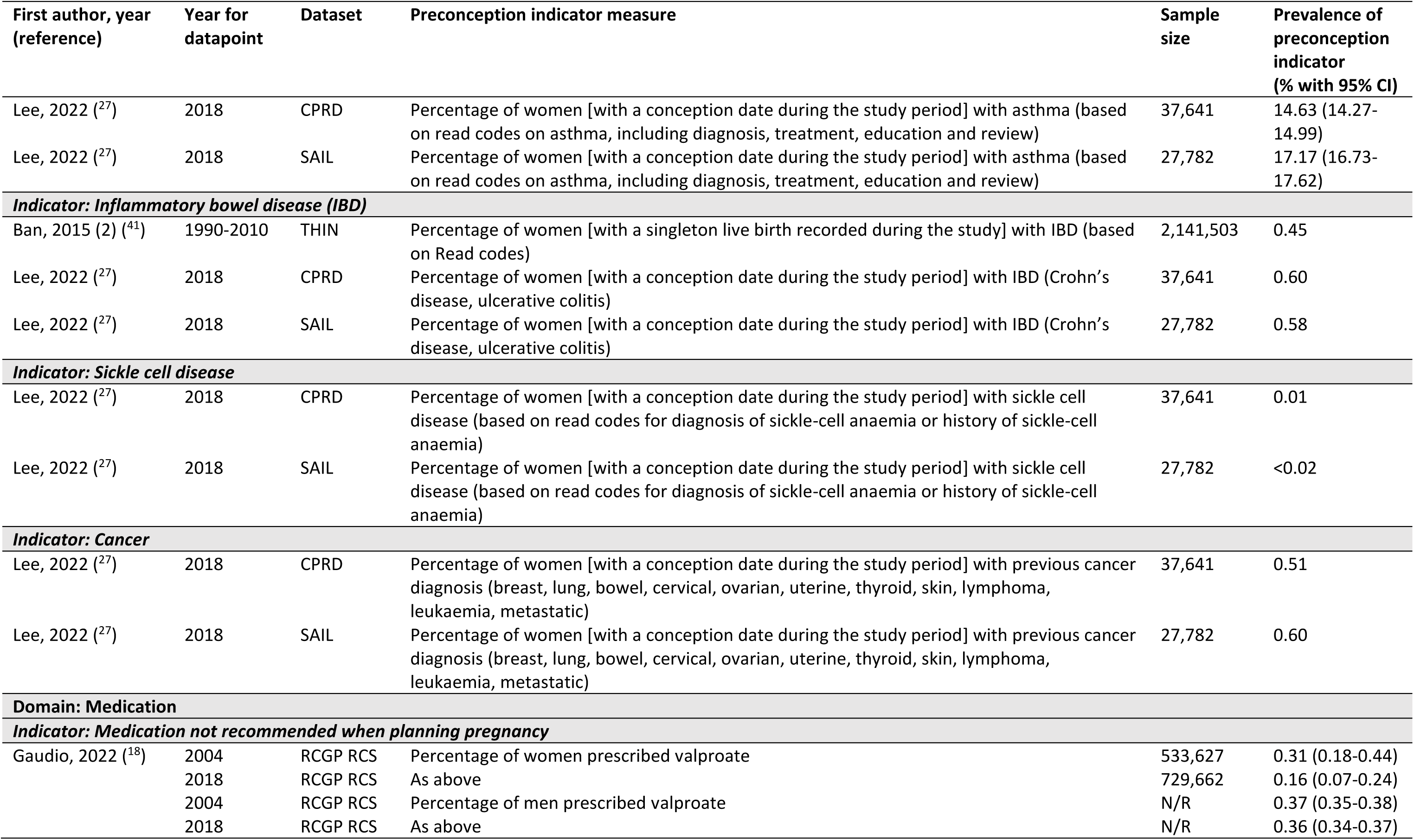
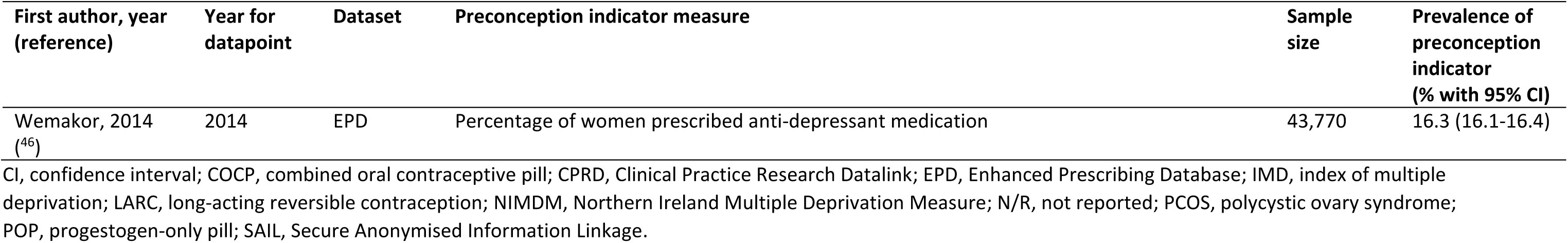
Prevalence of (and trends in) preconception indicators reported for people of reproductive age in UK routine primary care data.

The prevalence of preconception indicators reported across studies and primary care databases varied widely, possibly due to differences in preconception indicator definitions, year of data collection (Table 3), and study populations (Supplementary Table 5). The prevalence of preconception indicators defined in line with our scoping review (i.e. excluding individual methods of contraception, and prescribed folic acid supplements),^9^ ranged from 0.01% for sickle cell disease to >20% for each of advanced maternal age, previous caesarean section (among those with a recorded pregnancy), overweight, obesity, smoking and diagnosis of depression and anxiety among female patients (irrespective of pregnancy). Only three studies reported preconception indicators for male patients, showing for example that the prevalence of depression among fathers (9.2%) was lower compared with mothers (22.2%),^17^ and the proportion of patients prescribed valproate was comparable among female (0.31%) and male patients (0.37%) in 2004, but much lower among females (0.16%) than males (0.36%) in 2018.^18^

### Associations of preconception indicators with maternal and offspring outcomes

Five studies reported associations of preconception indicators (contraception prescription [n=1], sexually transmitted disease [n=1] and polycystic ovary syndrome [PCOS] [n=3]) with pregnancy and birth outcomes (Table 4). Outcome data were obtained from primary care data and/or linked HES data. Where two studies reported on comparable indicators and outcomes, consistent findings were shown for associations of PCOS with preterm delivery (<37 weeks gestational age) (positive association), high birthweight (>4kg) (no association) and low birthweight (<2.5kg) (inconclusive findings).^19, 20^

**Table 4.**
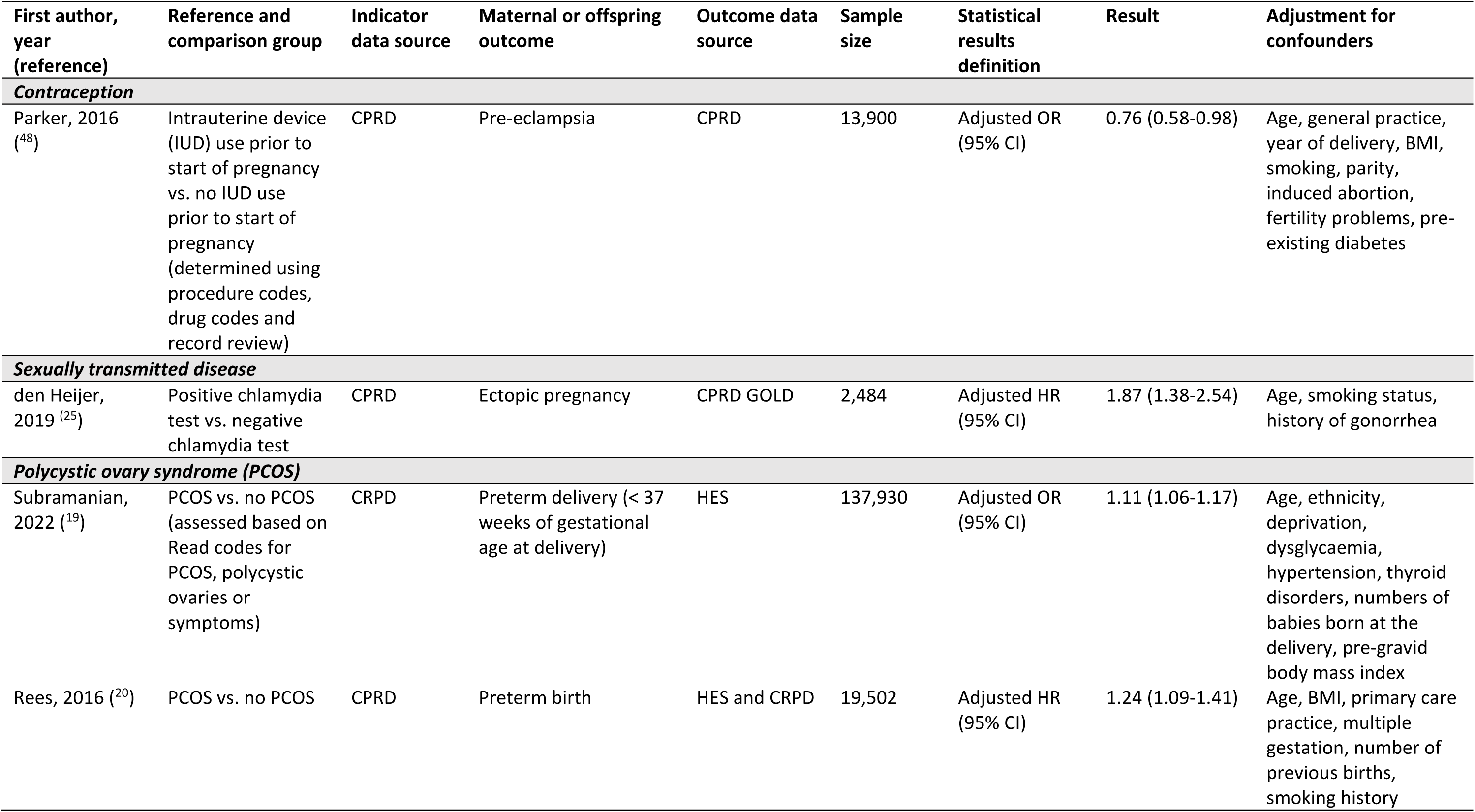
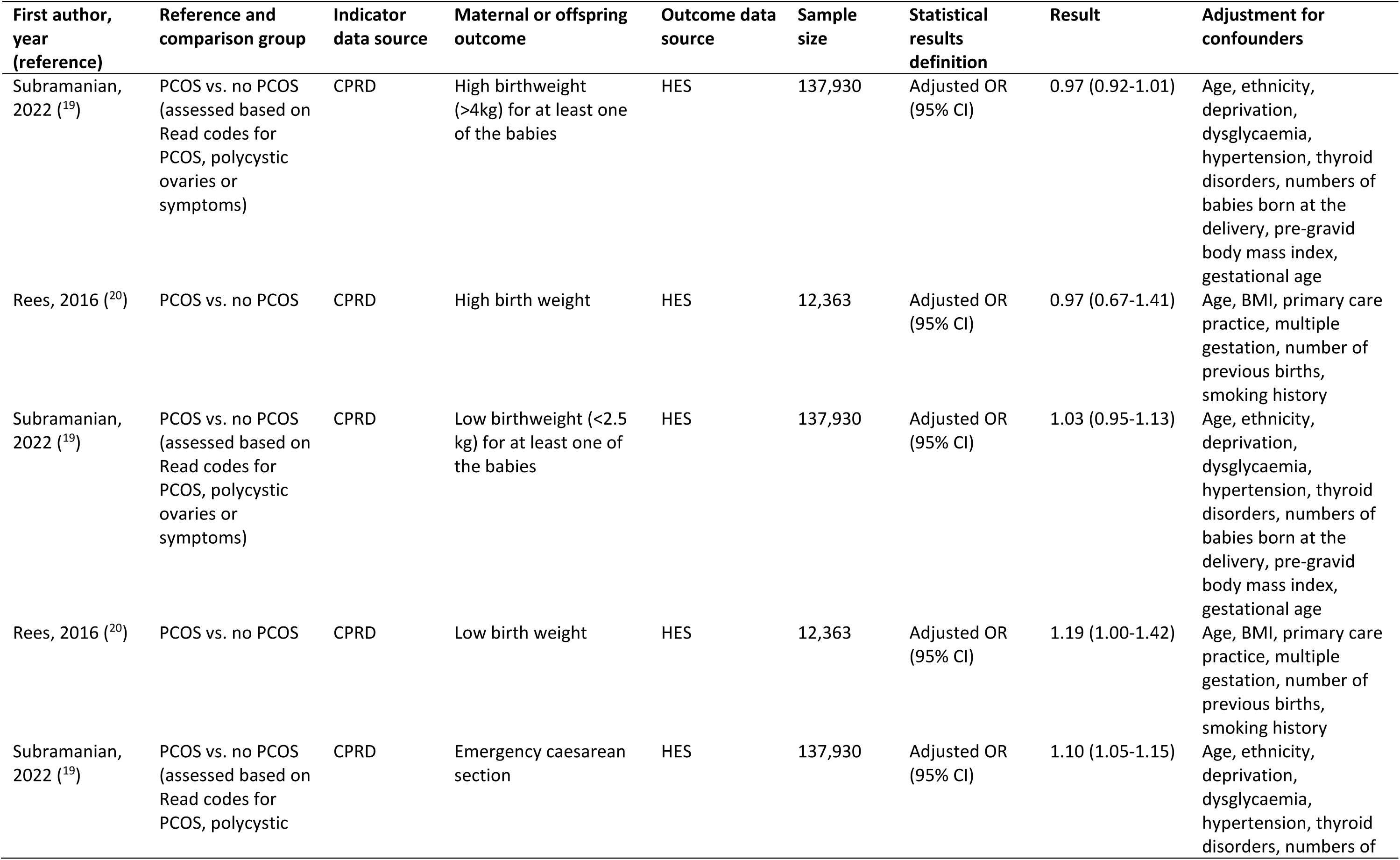
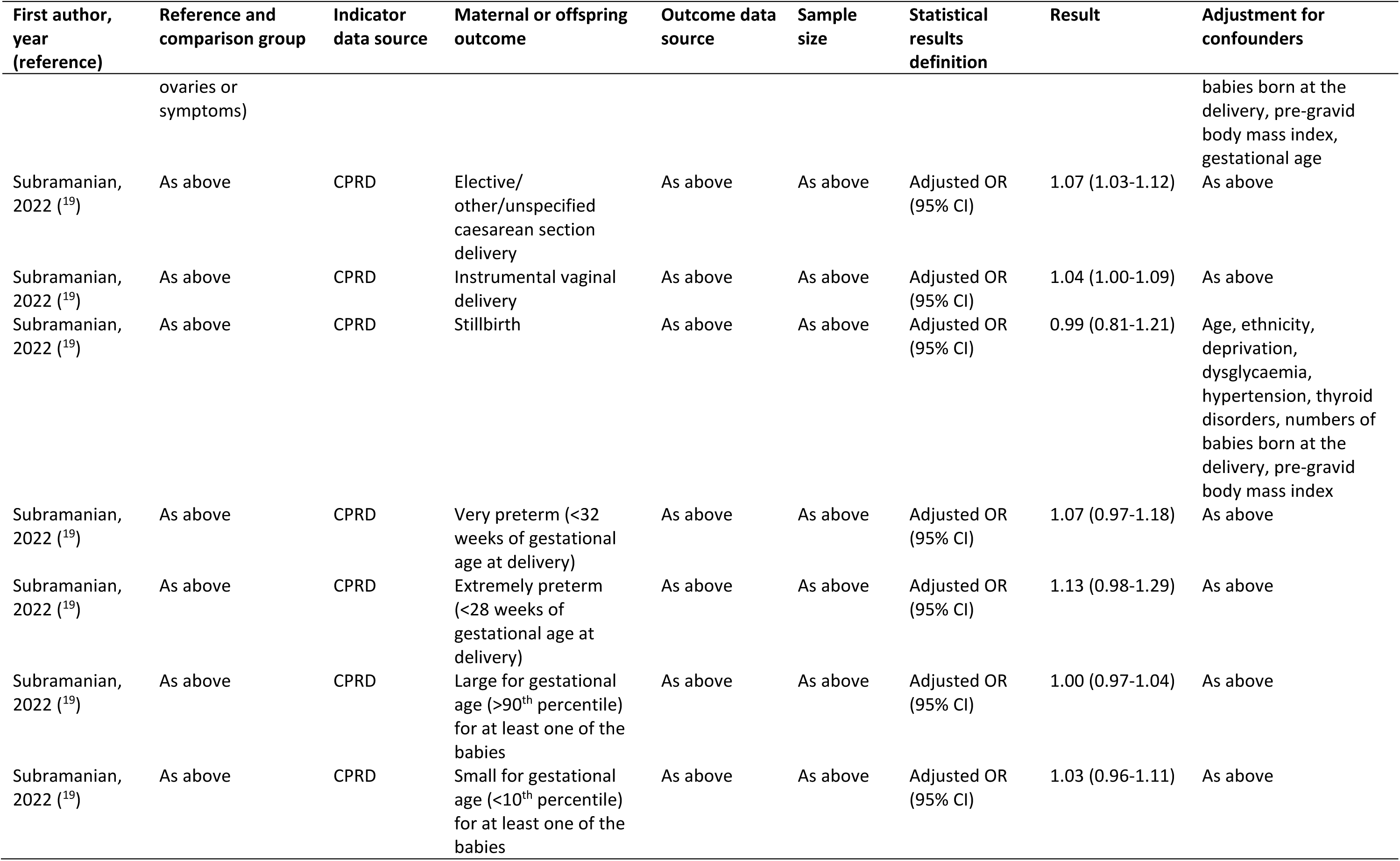
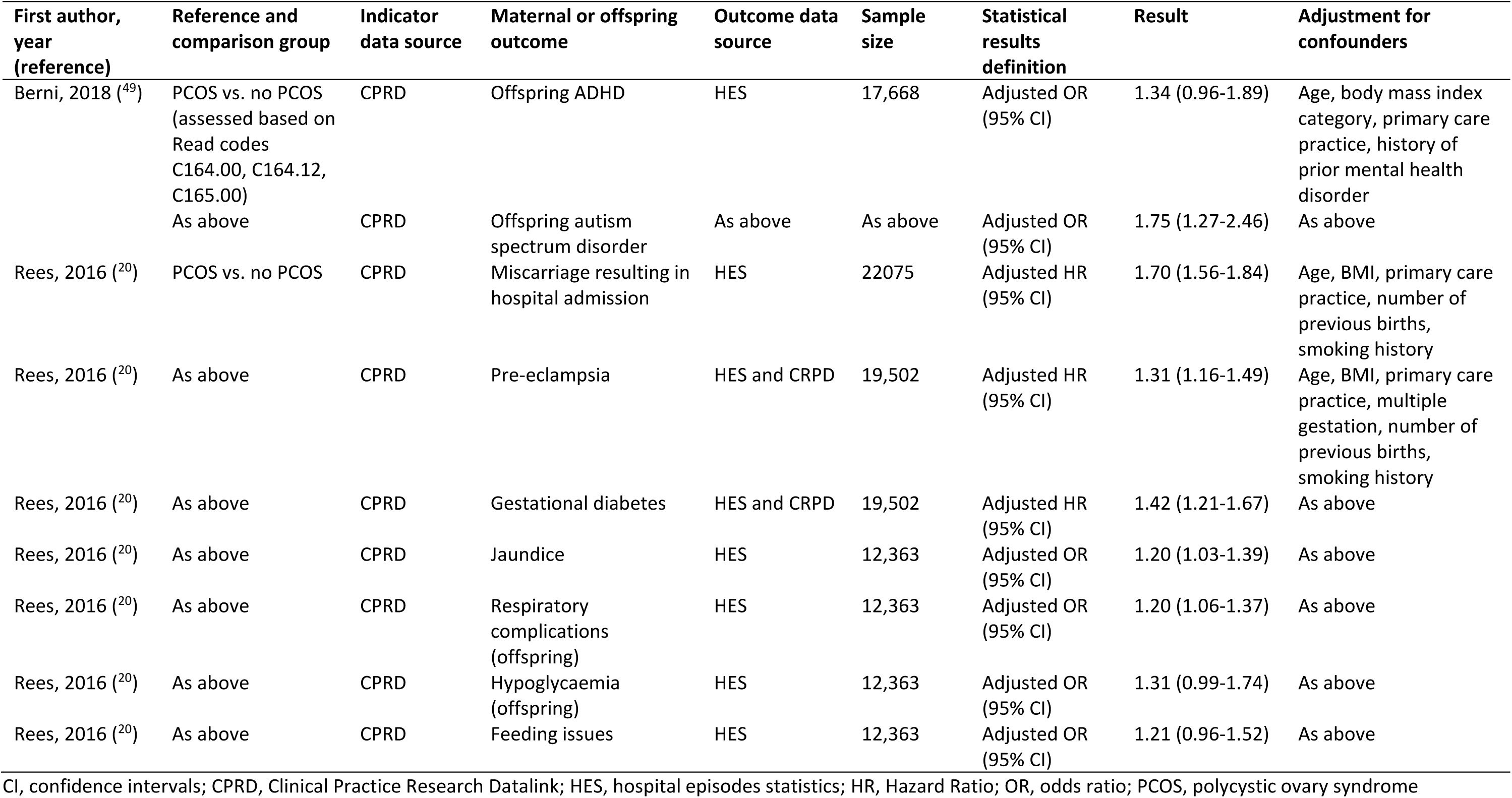
Associations of preconception indicators with outcomes in women and offspring.

| First author, year (reference) | Reference and comparison group | Indicator data source | Maternal or offspring outcome | Outcome data source | Sample size | Statistical results definition | Result | Adjustment for confounders |
| --- | --- | --- | --- | --- | --- | --- | --- | --- |
| **Contraception** |  |  |  |  |  |  |  |  |
| Parker, 2016 <sup>(48)</sup> | Intrauterine device (IUD) use prior to start of pregnancy vs. no IUD use prior to start of pregnancy (determined using procedure codes, drug codes and record review) | CPRD | Pre-eclampsia | CPRD | 13,900 | Adjusted OR (95% CI) | 0.76 (0.58-0.98) | Age, general practice, year of delivery, BMI, smoking, parity, induced abortion, fertility problems, pre-existing diabetes |
| **Sexually transmitted disease** |  |  |  |  |  |  |  |  |
| den Heijer, 2019 <sup>(25)</sup> | Positive chlamydia test vs. negative chlamydia test | CPRD | Ectopic pregnancy | CPRD GOLD | 2,484 | Adjusted HR (95% CI) | 1.87 (1.38-2.54) | Age, smoking status, history of gonorrhea |
| **Polycystic ovary syndrome (PCOS)** |  |  |  |  |  |  |  |  |
| Subramanian, 2022 <sup>(19)</sup> | PCOS vs. no PCOS (assessed based on Read codes for PCOS, polycystic ovaries or symptoms) | CRPD | Preterm delivery (< 37 weeks of gestational age at delivery) | HES | 137,930 | Adjusted OR (95% CI) | 1.11 (1.06-1.17) | Age, ethnicity, deprivation, dysglycaemia, hypertension, thyroid disorders, numbers of babies born at the delivery, pre-gravid body mass index |
| Rees, 2016 <sup>(20)</sup> | PCOS vs. no PCOS | CPRD | Preterm birth | HES and CRPD | 19,502 | Adjusted HR (95% CI) | 1.24 (1.09-1.41) | Age, BMI, primary care practice, multiple gestation, number of previous births, smoking history |
| Subramanian, 2022 <sup>(19)</sup> | PCOS vs. no PCOS (assessed based on Read codes for PCOS, polycystic ovaries or symptoms) | CPRD | High birthweight (>4kg) for at least one of the babies | HES | 137,930 | Adjusted OR (95% CI) | 0.97 (0.92-1.01) | Age, ethnicity, deprivation, dysglycaemia, hypertension, thyroid disorders, numbers of babies born at the delivery, pre-gravid body mass index, gestational age |
| Rees, 2016 <sup>(20)</sup> | PCOS vs. no PCOS | CPRD | High birth weight | HES | 12,363 | Adjusted OR (95% CI) | 0.97 (0.67-1.41) | Age, BMI, primary care practice, multiple gestation, number of previous births, smoking history |
| Subramanian, 2022 <sup>(19)</sup> | PCOS vs. no PCOS (assessed based on Read codes for PCOS, polycystic ovaries or symptoms) | CPRD | Low birthweight (<2.5 kg) for at least one of the babies | HES | 137,930 | Adjusted OR (95% CI) | 1.03 (0.95-1.13) | Age, ethnicity, deprivation, dysglycaemia, hypertension, thyroid disorders, numbers of babies born at the delivery, pre-gravid body mass index, gestational age |
| Rees, 2016 <sup>(20)</sup> | PCOS vs. no PCOS | CPRD | Low birth weight | HES | 12,363 | Adjusted OR (95% CI) | 1.19 (1.00-1.42) | Age, BMI, primary care practice, multiple gestation, number of previous births, smoking history |
| Subramanian, 2022 <sup>(19)</sup> | PCOS vs. no PCOS (assessed based on Read codes for PCOS, polycystic ovaries or symptoms) | CPRD | Emergency caesarean section | HES | 137,930 | Adjusted OR (95% CI) | 1.10 (1.05-1.15) | Age, ethnicity, deprivation, dysglycaemia, hypertension, thyroid disorders, numbers of babies born at the delivery, pre-gravid body mass index, gestational age |
| Subramanian, 2022 <sup>(19)</sup> | As above | CPRD | Elective/ other/unspecified caesarean section delivery | As above | As above | Adjusted OR (95% CI) | 1.07 (1.03-1.12) | As above |
| Subramanian, 2022 <sup>(19)</sup> | As above | CPRD | Instrumental vaginal delivery | As above | As above | Adjusted OR (95% CI) | 1.04 (1.00-1.09) | As above |
| Subramanian, 2022 <sup>(19)</sup> | As above | CPRD | Stillbirth | As above | As above | Adjusted OR (95% CI) | 0.99 (0.81-1.21) | Age, ethnicity, deprivation, dysglycaemia, hypertension, thyroid disorders, numbers of babies born at the delivery, pre-gravid body mass index |
| Subramanian, 2022 <sup>(19)</sup> | As above | CPRD | Very preterm (<32 weeks of gestational age at delivery) | As above | As above | Adjusted OR (95% CI) | 1.07 (0.97-1.18) | As above |
| Subramanian, 2022 <sup>(19)</sup> | As above | CPRD | Extremely preterm (<28 weeks of gestational age at delivery) | As above | As above | Adjusted OR (95% CI) | 1.13 (0.98-1.29) | As above |
| Subramanian, 2022 <sup>(19)</sup> | As above | CPRD | Large for gestational age (>90<sup>th</sup> percentile) for at least one of the babies | As above | As above | Adjusted OR (95% CI) | 1.00 (0.97-1.04) | As above |
| Subramanian, 2022 <sup>(19)</sup> | As above | CPRD | Small for gestational age (<10<sup>th</sup> percentile) for at least one of the babies | As above | As above | Adjusted OR (95% CI) | 1.03 (0.96-1.11) | As above |
| Berni, 2018 <sup>(49)</sup> | PCOS vs. no PCOS (assessed based on Read codes C164.00, C164.12, C165.00) | CPRD | Offspring ADHD | HES | 17,668 | Adjusted OR (95% CI) | 1.34 (0.96-1.89) | Age, body mass index category, primary care practice, history of prior mental health disorder |
|  | As above | CPRD | Offspring autism spectrum disorder | As above | As above | Adjusted OR (95% CI) | 1.75 (1.27-2.46) | As above |
| Rees, 2016 <sup>(20)</sup> | PCOS vs. no PCOS | CPRD | Miscarriage resulting in hospital admission | HES | 22075 | Adjusted HR (95% CI) | 1.70 (1.56-1.84) | Age, BMI, primary care practice, number of previous births, smoking history |
| Rees, 2016 <sup>(20)</sup> | As above | CPRD | Pre-eclampsia | HES and CRPD | 19,502 | Adjusted HR (95% CI) | 1.31 (1.16-1.49) | Age, BMI, primary care practice, multiple gestation, number of previous births, smoking history |
| Rees, 2016 <sup>(20)</sup> | As above | CPRD | Gestational diabetes | HES and CRPD | 19,502 | Adjusted HR (95% CI) | 1.42 (1.21-1.67) | As above |
| Rees, 2016 <sup>(20)</sup> | As above | CPRD | Jaundice | HES | 12,363 | Adjusted OR (95% CI) | 1.20 (1.03-1.39) | As above |
| Rees, 2016 <sup>(20)</sup> | As above | CPRD | Respiratory complications (offspring) | HES | 12,363 | Adjusted OR (95% CI) | 1.20 (1.06-1.37) | As above |
| Rees, 2016 <sup>(20)</sup> | As above | CPRD | Hypoglycaemia (offspring) | HES | 12,363 | Adjusted OR (95% CI) | 1.31 (0.99-1.74) | As above |
| Rees, 2016 <sup>(20)</sup> | As above | CPRD | Feeding issues | HES | 12,363 | Adjusted OR (95% CI) | 1.21 (0.96-1.52) | As above |

CI, confidence intervals; CPRD, Clinical Practice Research Datalink; HES, hospital episodes statistics; HR, Hazard Ratio; OR, odds ratio; PCOS, polycystic ovary syndrome

### Risk of bias and data quality

Risk of bias for findings on the prevalence of preconception indicators was generally low (n=18/21 studies), however, none of the studies received a minimal score (no bias) (Supplementary Table 2). Potential biases were introduced based on representativeness and sampling frame (e.g. excluding women with no pregnancy reported or no linked data available), and indicator definition and measurement (e.g. reporting individual methods of contraception rather than population prescribed contraception, or reliance on medication prescription rather than dispensing data). Moreover, details of non-response (e.g. impact of missing data) were not reported in approximately half the studies. Data quality for studies examining associations of preconception indicators with health outcomes was rated as good for four of the five studies (Supplementary Tables 3A-3B).

## Discussion

### Summary

This systematic review found that UK routine primary care data can provide valuable information on patients’ medical, behavioural and social risk factors before (a potential) pregnancy. Based on 42 included studies among people of reproductive age or women with a pregnancy recorded during the study period, the prevalence of 30 preconception indicators was reported. Findings showed that >20% of female patients of reproductive age would benefit from support with smoking cessation, and management of weight, depression and anxiety. This would optimise their own health, and improve their chance of a successful pregnancy and healthy baby if that is something they want. Limited research has used primary care data to examine preconception indicators among male patients, or associations of preconception indicators with pregnancy outcomes and longer-term maternal and offspring health outcomes.

### Strengths and limitations

This is the first systematic review to demonstrate how national routine primary care databases can be used to describe the population’s preconception health, to inform clinical practice and future research directions. Comprehensive, prospectively registered review methods were used. Our search was limited to UK primary care data and findings may not be generalisable to other countries. Preconception indicators were selected based on our previous scoping review;^9^ so potentially relevant indicators not included in this review or not reported in the included studies would have been missed. Moreover, some preconception indicators (such as dietary intake and physical activity) are not routinely recorded in general practice.

### Comparison with existing literature

Findings from our review complement our previous preconception report card based on the MSDS,^10^ showing that national routine health data are a valuable resource to describe and monitor women’s preconception health. Half of the preconception indicators identified in this review were also reported in the MSDS, with comparable prevalence estimates for most indicators (e.g. teenage pregnancy, previous caesarean delivery, overweight, obesity), while other indicators may be underreported in primary care (e.g. over the counter folic acid supplementation) or in the MSDS (e.g. mental health conditions).^10^ Published primary care data reported an additional 15 indicators not included in the MSDS (e.g. fertility problems, contraception, relevant medical conditions, teratogenic medication use). Linkage of these (and other) national routine health datasets would enhance the quality of preconception report cards and surveillance (Box 1). Based on linkage of primary care and HES datasets, findings from our review (n=2 studies^19, 20^) confirm the previously reported association of PCOS with increased risk of preterm delivery.^21^

**Box 1.**
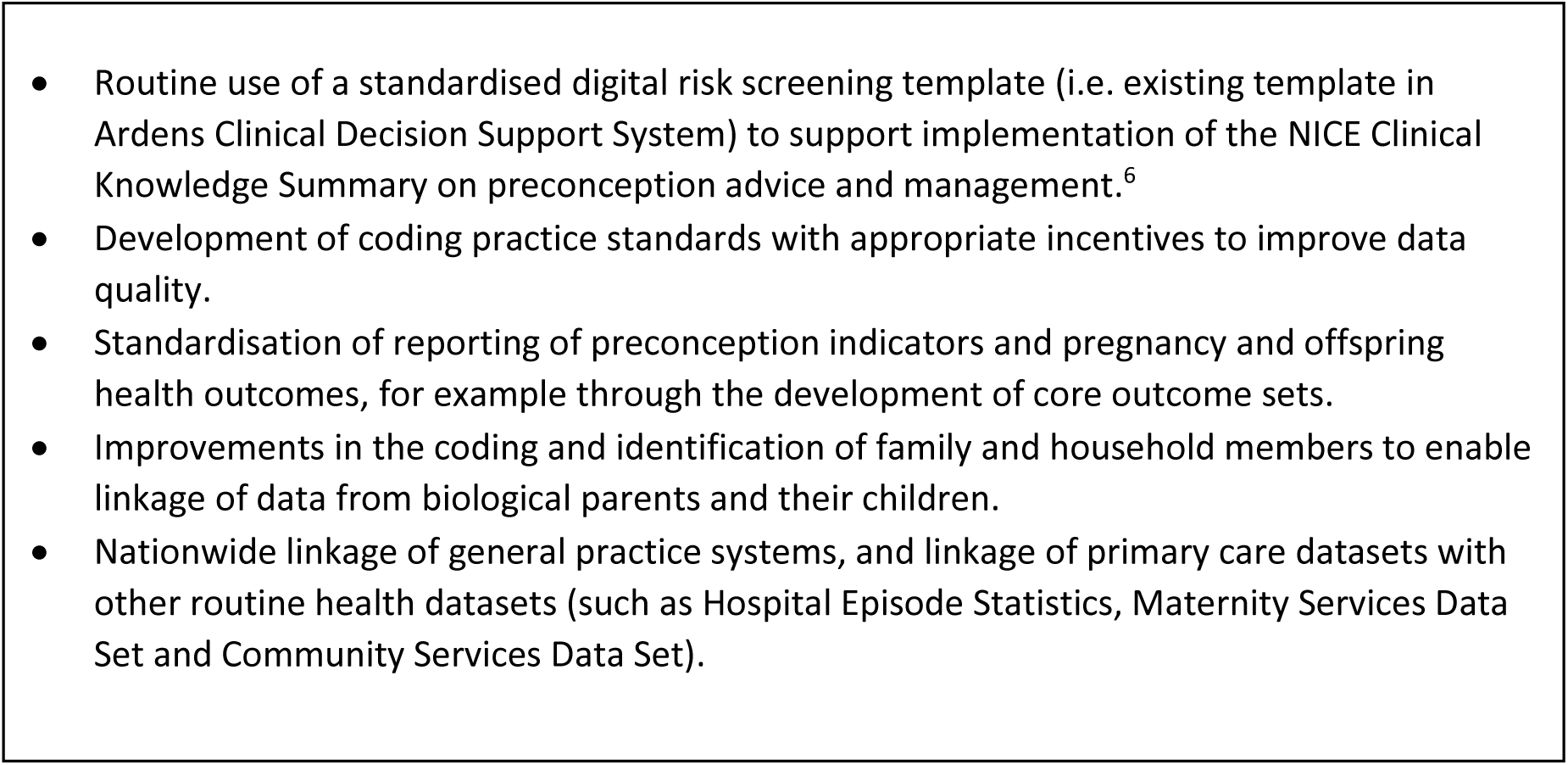
Recommendations to improve the use of UK routine primary care data for clinical practice, research and surveillance of preconception health and care.

Findings from our review are also in line with previous research reporting primary care data quality issues.^22–24^ Studies included in our review documented substantial missing data (20-60%) for ethnicity and BMI category, likely varying across sub-populations. Coding quality is related to financial incentives such as the Quality and Outcomes Framework (QOF), which may improve accurate recording of selected indicators but also distort prevalence estimates over time.^22^ The prevalence of some preconception indicators may be underestimated as not all conditions are solely diagnosed and coded in general practice (e.g. sexually transmitted disease),^25^ or medications and supplements prescribed (e.g. contraception, folic acid supplements).^26^ Another commonly reported limitation is the representation of selected general practices in research databases,^22, 23^ often limited to practices that use one of four main software platforms to manage electronic patient records (EPRs) and further determined by voluntary ‘opt ins’.^22, 23^ As a result, primary care databases may underrepresent specific regions and bias national prevalence estimates of preconception indicators and associations with health outcomes.

### Implications for research and clinical practice

Our findings demonstrate that many preconception indicators are routinely recorded in EPRs, allowing primary healthcare professionals to search for risk factors and provide individualised preconception care. A digital risk screening template has been developed in the Ardens Clinical Decision Support System based on the NICE Clinical Knowledge Summary,^6^ to support primary healthcare professionals to improve their preconception care practice, screening, coding and recording of indicators. Further work is required to co-develop practical guidance and resources to support integration of preconception care into every day clinical practice (Box 1).

Our findings identify the need to use standardised definitions when reporting preconception indicators (Box 1). Due to heterogeneity in definitions, the prevalence of preconception indicators across UK nations, and changes over time, could not be directly compared across studies. However, Lee and colleagues applied standardised definitions to CRPD (UK) and SAIL data (Wales), showing comparable prevalence estimates for some indicators (e.g. obesity, depression), but higher (e.g. smoking, underweight, anxiety, asthma) or lower (e.g. advanced maternal age) prevalence for other indicators, when comparing pregnant women in Wales with those in the UK overall.^27^ Moreover, standardised reporting within the same database showed, for example, increases over time in the prevalence of type 2 diabetes (1995-2017),^29^ alongside decreases in poor diabetes control (2004-2017).^29, 30^

Lastly, the limited reporting of male preconception indicators, and associations of preconception health with pregnancy, maternal and offspring health outcomes, calls for further research. Many of the preconception indicators reported for female patients are also relevant to male patients (e.g. smoking, obesity), with increasing evidence suggesting better paternal preconception health is associated with reduced risks of infertility and adverse pregnancy and offspring health and developmental outcomes.^31–33^ To enable further research, improvements are needed in the way that families (i.e. biological parents and their children) can be identified and data linked.^17, 34^ Primary care data also provide a unique opportunity to examine trajectories of preconception health during reproductive years irrespective of pregnancy, and to quantify the extent to which these reduce adverse pregnancy and offspring health outcomes. Future research would be enhanced by linkage of primary care and other routine health datasets beyond the identified existing linkages (e.g. MSDS and Community Services Data Set) to determine the short-and longer-term benefits of preconception care (Box 1).

## Conclusion

Routinely collected primary care data in the UK provide a valuable resource for research and surveillance, and can guide to provision of preconception care. Improvements in coding and reporting, and linkage of general practice systems and other national routine health datasets, would inform evidence-based provision of preconception care in primary care.

## Data Availability

All data produced in the present work are contained in the manuscript.

