## Supplementary file for "Preconception indicators and associations with health outcomes reported in UK routine primary care data: a systematic review"

**Table S1.** Search strategy used for each database Page 2

**Table S2.** Data quality: prevalence studies Page 4

**Table S3A.** Data quality: association studies (case-control studies) Page 7

**Table S3B.** Data quality: association studies (cohort studies) Page 7

**Table S4.** Characteristics of included studies reporting on the prevalence of preconception indicators in sub-population(s) Page 11

**Table S5.** In- and exclusion criteria of study populations Page 16

**References** Page 20

**Table S1.** Search strategy used for each database

| |
|---|
| **MEDLINE (Ovid)**<br>((Exp Preconception Care or Reproduction or Parents or (pre?conception or pre?conceptual or pre?pregnancy or reproductive* or conception or childbearing* or parent*).mp) and (Primary Health Care or General Practice or Family Practice or (primary care or general practice or GP or NHS or National Health Service).mp) and (UK or United Kingdom or Great Britain or England or Wales or Scotland or Northern Ireland or General Practice Research Database or General Practice Research Datalink or GPRD or Clinical Practice Research Database or Clinical Practice Research Datalink or CPRD or Health Improvement Network or THIN or QResearch or RCGP or National Diabets Audit or Secure Anonymised Information Linkage or SAIL or Enhanced Prescribing Database or Scottish Primary Care Information Resources or SPIRE or Primary Care Clinical Informatics Unit Research or PCCIUR or Welsh Longitudinal General Practice Dataset or WLGP or Optimum Patient Care Research Database or OPCRD).mp)) |
| **EMBASE (Ovid)**<br>((Exp Preconception Care or Reproduction or Parents or (pre?conception or pre?conceptual or pre?pregnancy or reproductive* or conception or childbearing* or parent*). ti,ab,tw) and (Primary Health Care or General Practice or Family Practice or (primary care or general practice or GP or NHS or National Health Service). ti,ab,tw) and (UK or United Kingdom or Great Britain or England or Wales or Scotland or Northern Ireland or General Practice Research Database or General Practice Research Datalink or GPRD or Clinical Practice Research Database or Clinical Practice Research Datalink or CPRD or Health Improvement Network or THIN or QResearch or RCGP or National Diabetes Audit or Secure Anonymised Information Linkage or SAIL or Enhanced Prescribing Database or Scottish Primary Care Information Resources or SPIRE or Primary Care Clinical Informatics Unit Research or PCCIUR or Welsh Longitudinal General Practice Dataset or WLGP or Optimum Patient Care Research Database or OPCRD).ti,ab,tw)) |
| **Scopus**<br>( TITLE-ABS-KEY ( pre?conception OR pre?conceptual OR pre?pregnancy OR reproductive* OR contraception OR childbearing* OR parent* ) AND TITLE-ABS-KEY ( "primary care" OR "general practice" OR gp OR nhs OR "National Health Service" ) AND TITLE-ABS-KEY ( ( uk OR "United Kingdom" OR "Great Britain" OR england OR wales OR scotland OR "Northern Ireland" OR "General Practice Research Database" OR "General Practice Research Datalink" OR GPRD OR "Clinical Practice Research Database" OR "Clinical Practice Research Datalink" OR CPRD OR "Health Improvement Network" OR thin OR QResearch OR RCGP OR "National Diabetes Audit" OR "Secure Anonymised Information Linkage" OR SAIL OR "Enhanced Prescribing Database" OR "Scottish Primary Care Information Resources" OR SPIRE OR "Primary Care Clinical Informatics Unit Research" OR PCCIUR OR "Welsh Longitudinal General Practice Dataset" OR WLGP OR "Optimum Patient Care Research Database" OR OPCRD) ) ) |
| **CINAHL**<br>((AB (pre?conception or pre?conceptual or pre?pregnancy or reproductive* or contraception or childbearing* or parent*)) and (AB ("primary care" or "general practice" or GP or NHS or "National Health Service")) and AB ((UK or "United Kingdom" or "Great Britain" or England or Wales or Scotland or "Northern Ireland" or "General Practice Research Database" or "General Practice Research Datalink" or GPRD or "Clinical Practice Research Database" or "Clinical Practice Research Datalink" or CPRD or "Health Improvement Network" or THIN or QResearch or RCGP or "National Diabetes Audit" or "Secure Anonymised Information Linkage" or SAIL or "Enhanced Prescribing Database" or "Scottish Primary Care Information Resources" or SPIRE or "Primary Care Clinical Informatics Unit Research" or PCCIUR or "Welsh Longitudinal General Practice Dataset" or WLGP or "Optimum Patient Care Research Database" or OPCRD)) or (TI (pre?conception or pre?conceptual or pre?pregnancy or reproductive* or contraception or childbearing* or parent*)) and (TI ("primary care" or "general practice" or GP or NHS or "National Health Service")) and TI ((UK or "United Kingdom" or "Great Britain" or England or Wales or Scotland or "Northern Ireland" |

| |
|---|
| or "General Practice Research Database" or "General Practice Research Datalink" or GPRD or "Clinical Practice Research Database" or "Clinical Practice Research Datalink" or CPRD or "Health Improvement Network" or THIN or QResearch or RCGP or "National Diabetes Audit" or "Secure Anonymised Information Linkage" or SAIL or "Enhanced Prescribing Database" or "Scottish Primary Care Information Resources" or SPIRE or "Primary Care Clinical Informatics Unit Research" or PCCIUR or "Welsh Longitudinal General Practice Dataset" or WLGP or "Optimum Patient Care Research Database" or OPCRD)) |
| **Web of Science**<br>(((TI=(pre?conception or pre?conceptual or pre?pregnancy or reproductive* or contraception or childbearing* or parent*)) OR (AB=(pre?conception or pre?conceptual or pre?pregnancy or reproductive* or contraception or childbearing* or parent*))) AND ((TI=("primary care" or "general practice" or GP or NHS or "National Health Service")) OR (AB=("primary care" or "general practice" or GP or NHS or "National Health Service"))) AND ((TI=(UK or "United Kingdom" or "Great Britain" or England or Wales or Scotland or "Northern Ireland" or "General Practice Research Database" or "General Practice Research Datalink" or GPRD or "Clinical Practice Research Database" or "Clinical Practice Research Datalink" or CPRD or "Health Improvement Network" or THIN or QResearch or RCGP or "National Diabetes Audit" or "Secure Anonymised Information Linkage" or SAIL or "Enhanced Prescribing Database" or "Scottish Primary Care Information Resources" or SPIRE or "Primary Care Clinical Informatics Unit Research" or PCCIUR or "Welsh Longitudinal General Practice Dataset" or WLGP or "Optimum Patient Care Research Database" or OPCRD)) OR (AB=(UK or "United Kingdom" or "Great Britain" or England or Wales or Scotland or "Northern Ireland" or "General Practice Research Database" or "General Practice Research Datalink" or GPRD or "Clinical Practice Research Database" or "Clinical Practice Research Datalink" or CPRD or "Health Improvement Network" or THIN or QResearch or RCGP or "National Diabetes Audit" or "Secure Anonymised Information Linkage" or SAIL or "Enhanced Prescribing Database" or "Scottish Primary Care Information Resources" or SPIRE or "Primary Care Clinical Informatics Unit Research" or PCCIUR or "Welsh Longitudinal General Practice Dataset" or WLGP or "Optimum Patient Care Research Database" or OPCRD)))) |

**Table S2.** Data quality: prevalence studies

| First author, year (reference) | Represen-tativeness | Sampling frame | Selection | Non-response | Data collection | Indicator definition | Reliable measurement | Similar measurement | Prevalence period | Appropriate numerator and denominator | Risk of bias rating |
|---|---|---|---|---|---|---|---|---|---|---|---|
| Ban, 2012 ([1]) | 1 | 1 | 0 | N/R | 0 | 0 | 0 | 0 | 0 | 0 | Low risk |
| Ban, 2015 (2) ([2]) | 0 | 0 | 0 | N/R | 0 | 0 | 1 | 0 | 0 | 0 | Low risk |
| Briggs, 2013 ([3]) | 0 | 0 | 0 | N/R | 0 | 1 | 1 | 0 | 0 | 0 | Low risk |
| Cea Soriano, 2013 ([4]) | 0 | 1 | 0 | N/R | 0 | 0 | 1 | 0 | 0 | 0 | Low risk |
| Cea Soriano, 2014 ([5]) | 0 | 1 | 0 | 0 | 0 | 0 | 1 | 0 | 0 | 0 | Low risk |
| Cea Soriano, 2018 ([6]) | 1 | 1 | 0 | N/R | 0 | 0 | 0 | 0 | 0 | 0 | Low risk |
| Coton, 2016 ([7]) | 1 | 1 | 0 | N/R | 0 | 0 | 0 | 0 | 0 | 0 | Low risk |
| Dave, 2010 ([8]) | 1 | 1 | 0 | N/R | 0 | 1 | 1 | 0 | 0 | 0 | Moderate risk |
| den Heijer, 2019 ([9]) | 0 | 0 | 0 | 1 | 0 | 1 | 1 | 0 | 0 | 0 | Low risk |
| Dhalwani, 2013 ([10]) | 0 | 0 | 0 | 0 | 0 | 0 | 1 | 0 | 0 | 0 | Low risk |
| Gaudio, 2021 ([11]) | 0 | 0 | 0 | 1 | 0 | 1 | 0 | 0 | 0 | 0 | Low risk |
| Gaudio, 2022 ([12]) | 0 | 0 | 0 | N/R | 0 | 1 | 1 | 0 | 0 | 0 | Low risk |
| Given, 2020 ([13]) | 0 | 0 | 0 | N/R | 0 | 0 | 1 | 0 | 0 | 0 | Low risk |
| Hope, 2022 ([14]) | 0 | 0 | 0 | 1 | 0 | 1 | 0 | 0 | 0 | 0 | Low risk |
| Lee, 2022 ([15]) | 1 | 1 | 0 | 1 | 0 | 1 | 0 | 0 | 0 | 0 | Moderate risk |
| Pasvol, 2022 ([16]) | 0 | 0 | 0 | 1 | 0 | 1 | 1 | 0 | 0 | 0 | Low risk |
| Rowlands, 2000 ([17]) | 0 | 0 | 0 | N/R | 0 | 1 | 1 | 0 | 0 | 0 | Low risk |
| Smith, 2020 ([18]) | 1 | 1 | 0 | 1 | 0 | 0 | 0 | 0 | 0 | 0 | Low risk |
| Subramanian, 2022 ([19]) | 1 | 1 | 1 | 1 | 0 | 0 | 1 | 0 | 0 | 0 | Moderate risk |
| Syed, 2022 ([20]) | 1 | 1 | 0 | 0 | 0 | 0 | 1 | 0 | 0 | 0 | Low risk |

| | | | | | | | | | | |
|---|---|---|---|---|---|---|---|---|---|---|
| Wemakor, 2014 ([21]) | 0 | 0 | 0 | 1 | 0 | 1 | 1 | 0 | 0 | 0 | Low risk |

N/R, not reported.

**Scoring guide:**
Risk of bias tool for population-based prevalence studies based on Hoy et al (ref)

**External validity**
1. Was the study's target population a close representation of the general population of people of reproductive age (in terms of socio-demographic characteristics)?
Yes = 0
No = 1

2. Was the sampling frame a true or close representation of the target population?
Yes = 0
No = 1

3. Was some form of random selection used to select the sample, or was a census undertaken?
Yes = 0
No = 1

4. Was the likelihood of non-response bias minimal?
Yes = 0 (*response rate was ≥80%, or an analysis showed no significant differences in socio-demographic characteristics between patients included and lost due to non-response/missing data*)
No = 1 (*response rate was <80%, or if analysis comparing included and excluded patients was done it showed a significant difference in socio-demographic characteristics*)

**Internal validity**
5. Were data collected directly from patients (as opposed to a proxy)?
Yes = 0 (*all data collected directly from patient*)
No = 1 (*some or all data collected from a proxy (e.g. parent, carer, linked non-patient level data)*)

6. Was an acceptable indicator definition used in the study?
Yes = 0 (*indicator definition in line with Schoenaker et al 2022 review*)
No = 1

7. Was a reliable and accepted method used to assess or measure the indicator?
Yes = 0
No = 1

8. Was the same mode of data collection used for all patients?
Yes = 0
No = 1

9. Was the length of the prevalence period for the indicator of interest appropriate? (*e.g. appropriate period to make sure the indicator is not under/overestimated*)
Yes = 0
No = 1

10. Were the numerator(s) and denominator(s) for the indicator of interest appropriate
Yes = 0 (*numerator = in line with indicator definition, denominator = all people of reproductive age in the dataset*)
No = 1

**Summary score:** 0-3: low risk; 4-6: moderate risk; 7-9: high risk

**Table S3A.** Data quality: association studies (case-control studies)

| | Selection | | | Comparability | Exposure | | | |
|---|---|---|---|---|---|---|---|---|
| **First author, year (reference)** | **Case definition** | **Representativeness** | **Selection of controls** | **Comparability** | **Exposure ascertainment** | **Same method for cases and controls** | **Non-response rate** | **Data quality rating** |
| Subramanian, 2022 ([19]) | A* | A* | A* | A* | A* | A* | D | Good quality |
| Bernie, 2018 ([22]) | A* | A* | A* | A*B* | A* | A* | D | Good quality |
| Parker, 2016 ([23]) | A* | A* | A* | A*B* | A* | A* | A* | Good quality |
| Rees, 2016 ([24]) | A* | A* | A* | A*B* | A* | A* | D | Good quality |

**Table S3B.** Data quality: association studies (cohort studies)

| | Selection | | | Comparability | Outcome | | | |
|---|---|---|---|---|---|---|---|---|
| **First author, year (reference)** | **Representativeness of exposed cohort** | **Selection of non-exposed cohort** | **Exposure ascertainment** | **Comparability** | **Outcome assessment** | **Follow-up duration** | **Follow-up rate** | **Data quality rating** |
| den Heijer, 2019 ([9]) | C | A* | A* | A*B* | A* | A* | D | Fair quality |

**Scoring guide:**

**Newcastle Ottawa Quality Assessment Scale – <u>Case control studies</u>**

Selection (select one option)
1) Is the case definition adequate?
a) yes, with independent validation (*e.g. >1 person has validated data, or data from primary record source such as clinical record*) *
b) yes (*e.g. record linkage or based on self-reports with no reference to primary record*)
c) no description

2) Representativeness of the cases
a) consecutive or obviously representative series of cases (*e.g. all cases with outcome of interest over a period of time, all cases in a defined clinic or area, random sample*) *
b) potential for selection biases or not stated

3) Selection of Controls
a) community controls (e.*g. same community as cases*) *
b) hospital controls (*e.g. same community as cases but derived from hospitalised population*)
c) no description

Comparability (select one or both options)
4) Comparability of cases and controls on the basis of the design or analysis
a) study controls for age (*controlled through matching of cases and controls, or confounders are adjusted for in the analysis*) *
b) study controls for any additional factor (*e.g. ethnicity, SES, previous pregnancy, BMI*) *

Exposure (select one option)
5) Ascertainment of exposure
a) secure record (*e.g. primary care record*) *
b) structured interview where blind to case/control status *
c) interview not blinded to case/control status
d) written self-report
e) no description

6) Same method of ascertainment for cases and controls
a) yes *
b) no

7) Non-Response rate
a) same rate for both groups *
b) non respondents described
c) rate different and no designation
d) not reported or unclear

**Newcastle Ottawa Quality Assessment Scale – Cohort studies**

Selection (select one option)
1) Representativeness of the exposed cohort
a) truly representative of the average population of people of reproductive age in the community *
b) somewhat representative of the average population of people of reproductive age in the community *
c) selected group of people not representative of the average population of people of reproductive age in the community
d) no description of the derivation of the cohort

2) Selection of the non-exposed cohort
a) drawn from the same community (e.g. same data source) as the exposed cohort *
b) drawn from a different source
c) no description of the derivation of the non-exposed cohort

3) Ascertainment of exposure
a) secure record (*e.g. primary care record*) *
b) structured interview *
c) written self-report
d) no description

Comparability (select one or both options)
4) Comparability of cohorts on the basis of the design or analysis
a) study controls for age (*e.g. through confounder adjustment in the analysis*) *
b) study controls for any additional factor (*e.g. ethnicity, SES, previous pregnancy, BMI*) *

Outcome (select one option)
5) Assessment of outcome
a) independent or blind assessment, or confirmation of the outcome by reference to secure records (*e.g. medical records*) *
b) record linkage *
c) self report
d) no description

6) Was follow-up long enough for outcomes to occur
a) yes (*e.g. follow up throughout pregnancy for pregnancy complications, follow up throughout childhood for childhood outcomes*) *
b) no

7) Adequacy of follow up of cohorts
a) complete follow up - all subjects accounted for *
b) subjects lost to follow up unlikely to introduce bias (*e.g. response rate was ≥80%, or an analysis showed no significant differences in demographic characteristics between patients included and lost due to non-response/missing data, or multiple imputation use*) *
c) follow up rate <80% and no description of those lost
d) no statement

**Data quality rating:**
- Good quality: 3 stars in selection domain & 1 or 2 stars in comparability domain & 2 or 3 stars in outcome/exposure domain

- Fair quality: 2 stars in selection domain & 1 or 2 stars in comparability domain & 2 or 3 stars in outcome/exposure domain
- Poor quality: 0 or 1 star in selection domain or 0 stars in comparability domain or 0 or 1 stars in outcome/exposure domain

**Table S4.** Characteristics of included studies reporting on the prevalence of preconception indicators in sub-population(s)

| First author, year (reference) | Dataset | Country | Study design | Data collection period | Total or maximum sample size | Population characteristics: sex, age | Preconception indicators reported | Maternal and offspring outcomes reported |
|---|---|---|---|---|---|---|---|---|
| ***Clinical Practice Research Datalink (CPRD)*** | | | | | | | | |
| Haase, 2023 ([25]) | CPRD GOLD | UK | Cohort study | 1987-2021 | 9,995 | Female, 18-45 years | Deprivation, weight, smoking, contraception, diabetes mellitus, hypertension, asthma | None |
| Channon, 2022 ([26]) | CPRD GOLD | UK | Cohort study | 2009-2018 | 318,040 | Female, 16-48 years | Contraception, folic acid prescription, weight | None |
| Hope, 2022 ([14]) | CPRD GOLD | UK | Cohort study | 1990-2017 | 2,680,149 | Female, 14-45 years | Deprivation, contraception, smoking, STDs, PCOS, endometriosis, cancer, cervical screening | None |
| Subramanian, 2022 ([19]) | CPRD GOLD linked with HES | England | Cohort and case-control study | 1997-2020 | 299,866 | Female, 15-49 years | Maternal age, deprivation, weight | Preterm delivery, mode of delivery, high or low birthweight, stillbirth, small and large for gestational age |
| Syed, 2022 ([20]) | CPRD GOLD linked with HES and Office for National Statistics (ONS) mortality register | England | Cohort study | 2004- 2018 | 211,393 | Female, 16-55 years | No prevalence data reported: housing, alcohol consumption, substance use, eating disorder, depression, anxiety, domestic abuse | None |
| Ma, 2020 ([27]) | N/A | England, Wales, Scotland | Cohort study | 2004-2014 | 3,281,667 | Female, 13-54 years | No prevalence data reported: contraception | None |
| Jackson, 2019 ([28]) | CPRD GOLD linked with HES and IMD | UK | Case-control study | 2011-2016 | 217,850 | Female, 15-49 years | Deprivation, domestic abuse, contraception, alcohol consumption, depression | None |

| First author, year (reference) | Dataset | Country | Study design | Data collection period | Total or maximum sample size | Population characteristics: sex, age | Preconception indicators reported | Maternal and offspring outcomes reported |
|---|---|---|---|---|---|---|---|---|
| Berni, 2018 ([22]) | CPRD GOLD linked with HES | UK | Case-control study | 2000-2014 | 16,355 cases and 16,355 controls | Female, mean age 27 years (SD 7) | Weight, smoking, alcohol, hypertension | Offspring ADHD and autism spectrum disorder |
| Richardson, 2018 ([29]) | CPRD linked with HES and IMD | UK | Case-control study | 2002-2012 | 44,260 | Female, 18-55 years | Smoking, alcohol | None |
| Parker, 2016 ([23]) | N/A | UK | Nested case-control study | 1993-2010 | 13,900 | Female, mean age 29 years (SD 6) | Maternal age, contraception, weight, smoking, fertility problems, pre-existing diabetes | Pre-eclampsia |
| Rees, 2016 ([24]) | CPRD linked with HES | UK | Nested case-control study | 2000-2012 | 27,204 | Female, 15-44 years | Maternal age, PCOS, routine GP check-up in past year, contraception, history of assisted reproduction, weight, hypertension, smoking | Miscarriage, pre-eclampsia, gestational diabetes, premature delivery, delivery method, jaundice, respiratory complications, feeding issues, high birth weight, low birth weight, hypoglycaemia |
| Nightingale, 2000 ([30]) | CPRD and UK MediPlus database (not linked) | UK | Case-control studies | 1992-1997 | 1,851 | Female, 15-49 years | Contraception, weight, smoking, asthma, hypertension, thromboembolism | None |
| ***General Practice Research Database (GPRD)*** | | | | | | | | |
| Briggs, 2013 ([3]) | N/A | UK | Repeated cross-sectional study | 2004-2010 | 1,103,669 | Female, 15-49 years | Weight, hypertension, smoking, thromboembolism, cardiovascular disease | None |

| First author, year (reference) | Dataset | Country | Study design | Data collection period | Total or maximum sample size | Population characteristics: sex, age | Preconception indicators reported | Maternal and offspring outcomes reported |
|---|---|---|---|---|---|---|---|---|
| Khashan, 2012 ([31]) | N/A | UK | Cohort study | 1990-2008 | 100,000 | Female, 13-50 years | Maternal age, weight, deprivation, smoking, alcohol, diabetes, hypertension, asthma, inflammatory bowel disease | Not examined in relation with preconception indicator: miscarriage, ectopic pregnancy, pre-eclampsia, stillbirth |
| Shawe, 2008 ([32]) | N/A | UK | Case-control study | 2001 | 9,462 | Female, 15-44 years | Contraception | None |
| Howard, 2004 ([33]) | N/A | UK | Case-control study | 1996-1998 | 986 | Female, 15-44 years | Severe mental health conditions, substance use, prescribed medication, smoking, alcohol Questionnaire data: social factors | None |
| Seaman, 2003 ([34]) | N/A | UK | Repeated cross-sectional study | 1992-1998 | N/R | Female, 15-39 years | Contraception, PCOS | None |
| Shorvon, 2002 ([35]) | N/A | UK | Cross-sectional study | 1995 | 2,341 | Female, 15-45 years | Contraception | None |
| ***The Health Improvement Network (THIN)*** | | | | | | | | |
| Cea Soriano, 2018 ([6]) | N/A | UK | Cohort study and case-control study | 1995-2012 | 251,581 | Female, 15-45 years | Maternal age, smoking, weight, depression, anxiety, asthma, epilepsy, hypertension, PCOS | None |
| Cea Soriano, 2016 ([36]) | N/A | UK | Cohort study | 2002-2010 | 22841 | Female, 12-49 years | Contraception, PCOS, fertility problems | None |
| Coton, 2016 ([7]) | N/A | UK | Cohort study | 1995-2012 | 301,794 | Female, 16 and over | Deprivation, maternal age, smoking, weight, hypertension, glycaemic control | None |

| First author, year (reference) | Dataset | Country | Study design | Data collection period | Total or maximum sample size | Population characteristics: sex, age | Preconception indicators reported | Maternal and offspring outcomes reported |
|---|---|---|---|---|---|---|---|---|
| Ban, 2015 (1) ([37]) | N/A | UK | Cohort study | 1990-2013 | 1,259 | Female, 15-44 years | Folic acid supplementation | None |
| Ban, 2015 (2) ([2]) | N/A | UK | Cohort study | 1990-2010 | 2,141,503 | Female, 15-44 years | Deprivation, contraception | None |
| Cea Soriano, 2014 ([5]) | N/A | UK | Cohort study | 2004-2010 | N/R | Female, 18-44 years | Previous pregnancy loss | None |
| Dave, 2010 ([8]) | Use of family identification number to link mothers, fathers and children living in the same household | UK | Cohort study | 1993-2007 | 86,957 | Female and male, 15-≥35 years | No prevalence data reported: maternal and paternal age, deprivation | None |
| ***Royal College of General Practitioners (RCGP) Research and Surveillance Centre (RSC) network*** | | | | | | | | |
| Gaudio, 2022 ([12]) | N/A | England | Repeated cross-sectional study | 2004-2018 | 729,662 | Female and male, 12-46 years | Deprivation, smoking, epilepsy, mental health condition, contraception, folic acid supplementation | None |
| Gaudio, 2021 ([11]) | N/A | England | Repeated cross-sectional study | 2004-2017 | 465,898 | Female, 16-45 years | Deprivation, smoking, contraception, weight, hypertension, medication not recommended in pregnancy | None |
| ***Scottish Programme for Improving Clinical Effectiveness in Primary Care (SPICE)*** | | | | | | | | |
| Krishnamoorthy, 2008 ([38]) | N/A | Scotland | Repeated cross-sectional study | 2000-2006 | N/R | Female, 10-19 years | Contraception | None |
| Krishnamoorthy, 2005 ([39]) | SPICE database combined with anonymous data from family planning clinics | Scotland | Cross-sectional study | 1999-2000 | 35,414 | Female, 10-16 years | Contraception | None |
| ***Other datasets*** | | | | | | | | |
| Nwaru, 2021 ([40]) | Optimum Patient Care Research Database | UK | Cohort study | 2000-2016 | 83,084 | Female, 16-45 years | Deprivation, asthma, contraception, weight, smoking | None |

| First author, year (reference) | Dataset | Country | Study design | Data collection period | Total or maximum sample size | Population characteristics: sex, age | Preconception indicators reported | Maternal and offspring outcomes reported |
|---|---|---|---|---|---|---|---|---|
| Smith, 2021 ([41]) | Electronic primary care records for ALSPAC study participants linked with ALSPAC questionnaire data | England | Prospective cohort study | 1991-2014 | 11,807 | Female and male, 15-23 years | Depression, anxiety<br>Questionnaire data: maternal age, housing, education | None |
| Reddy, 2014 ([42]) | Computerised data from general practices throughout Scotland collected by the Primary Care Clinical Informatics Unit (PCCIUR) of the University of Aberdeen | Scotland | Repeated cross-sectional study | 2004-2009 | ~340,000 in each year | Female, 12-55 years | No prevalence data reported: contraception, smoking, deprivation | None |

ALSPAC, Avon Longitudinal Study of Parents and Children; CPRD, Clinical Practice Research Datalink; EPD, Enhanced Prescribing Database; HES, Hospital Episodes Statistics; IMD, index of multiple deprivation; PCOS, polycystic ovary syndrome; SD, standard deviation.

**Table S5.** In- and exclusion criteria of study populations

| First author, year (reference) | Study population description |
|---|---|
| Ban, 2012 ([1]) | Women aged 15-45 years who were registered with a general practice and who had at least one recorded pregnancy ending in a live birth during the study period (446 general practices). |
| Ban, 2015 (1) ([37]) | Women aged 15-44 years who were registered with a general practice and who had a singleton live birth during the study period.<br>Exclusion criteria: women registered for less than three months before conception or no precise record of gestational week or an expected due date, women with a diagnosis of diabetes or prescription for anti-diabetic medication before or during pregnancy. |
| Ban, 2015 (2) ([2]) | Women aged 15-44 years who were registered with a general practice. |
| Berni, 2018 ([22]) | Women aged 14 or above who were registered with a general practice and had a pregnancy recorded during the study period and data linked with Hospital Episode Statistics.<br>Cases: women with polycystic ovary syndrome (PCOS).<br>Controls: women with no history of PCOS. |
| Briggs, 2013 ([3]) | Women aged 15-49 years who were registered with a general practice and had at least one prescription for a combined hormonal contraceptive and at least one UK Medical Eligibility Criteria for Contraceptive Use (UKMEC) Category 3 or 4 risk factor. |
| Cea-Soriano, 2013 ([4]) | Women aged 12-49 years who were registered with a general practice for at least 5 years and had a prescription history of at least 1 year (562 general practices [6.2% of the UK population]). |
| Cea-Soriano, 2014 ([5]) | Women aged 18-44 years who were registered with a general practice for at least 5 years and had a prescription history of at least 1 year. |
| Cea-Soriano, 2016 ([36]) | Women aged 12-49 years who were registered with a general practice for at least 5 years and had a prescription history of at least 1 year, who were new users of cyproterone acetate/ethinylestradiol (CPA/EE), levonorgestrel/EE (LNG/EE) or drospirenone/EE (DRSP/EE). (578 general practices [6% of the UK population]). |
| Cea-Soriano, 2018 ([6]) | Women aged 15-45 who were registered with a general practice for at least 1 year, who had type 1 or type 2 diabetes before pregnancy and a pregnancy recorded during the study period. |
| Channon, 2022 ([26]) | Women aged 16-48 years who were registered with a general practice and had at least one consultation identified as LARC-related during the study period. |
| Coton, 2016 ([7]) | Pregnant women aged 16 and over who were registered with a general practice and give birth during the study period. General practices were excluded if they did not meet acceptable computer use (ACU) or acceptable mortality rates (AMR) data quality measures. |
| Dave, 2010 ([8]) | Women with a child and/or pregnancy living in a household with a single adult man who had registered with the practice before the child was aged 1 year (putative father). |
| den Heijer, 2019 ([9]) | Women aged 12-25 years who were registered with a general practice and had data linked to the English Index for Multiple Deprivation. Women with a history of hysterectomy, bilateral oophorectomy/ovariectomy or sterilisation were excluded. |
| Dhalwani, 2013 ([10]) | Women aged 15-49 years who were registered with a general practice and who contributed one or more years of active registration during the study period (495 general practices). |
| Gaudio, 2021 ([11]) | Women aged 16-45 years who were registered with a general practice and had a diagnosis of type 1 or type 2 diabetes. |

| First author, year (reference) | Study population description |
|---|---|
| | Exclusion criteria: women who were pregnant during the study period (excluded for the duration of pregnancy); women with menopause (excluded from the date of the documented menopause). |
| Gaudio, 2022 ([12]) | Women and men aged 12-46 years who were registered with a general practice at any point and for a complete year during the study period. |
| Given, 2020 ([13]) | Women aged 12-49 years who were registered with a general practice. |
| Haase, 2023 ([25]) | Women aged 18-45 years with a diagnosis of PCOS and a BMI of ≥18.5 kg/$m^2$ (study 1) or a BMI of ≥25 kg/$m^2$ (study 2).<br>Exclusion criteria: women with an intrauterine contraceptive device, cancer, pregnancy or thyroid disorder during the baseline period, or a prescription for contraceptives or ovulation induction drugs at the index date (time of PCOS diagnosis). |
| Hope, 2022 ([14]) | Women aged 14-45 years with at least once pregnancy during the study period who were registered for at least two years with a general practice. |
| Howard, 2004 ([33]) | Women aged 15-44 years who were registered with a general practice.<br>Cases: women with a diagnosis of psychotic disorder at any time up to the index birth.<br>Controls: women with no history of psychotic disorder who had children during the same period. |
| Jackson, 2019 ([28]) | Women aged 15-49 years who were registered with a general practice.<br>Cases: women with at least one record of emergency contraception consultation within the study period.<br>Controls: women with no record of emergency contraception consultation within the study period. Controls were excluded if they had any indication that they would not have been eligible for, or needed, emergency contraception. |
| Krishnamoorthy, 2005 ([39]) | Women aged 10-16 years who were registered with a general practice (161 general practices). |
| Krishnamoorthy, 2008 ([38]) | Women aged 10-19 years who were registered with a general practice (31% of Scottish general practices). |
| Khashan, 2012 ([31]) | Pregnant women aged 13-50 years with up-to-standard data (i.e. prospective data recording by the general practice) who were selected by stratified random sampling from all women with a diagnosis of pregnancy in their clinical or referral records during the study period. |
| Lee, 2022 ([15]) | Pregnant women aged 15-49 years with a conception date during the study period, with data that met standard quality checks (940 general practices in CRPG GOLD [4% of UK general practices], SAIL [80% of Welsh general practices]). |
| Ma, 2020 ([27]) | Woman aged 13-54 who were registered with a general practice. Women became eligible when they turned 13 in any study year; women who turned 55 were censored, as were any who died, or if their practice stopped contributing data (over 600 general practices). |
| Nightingale, 2000 ([30]) | Women aged 15-49 years who were registered with a general practice who had evidence of idiopathic venous thromboembolism, were treated with oral anticoagulants, and exposed to combined oral contraceptives at the time of the event.<br>Exclusion criteria: pregnancy within 6 weeks prior to the event, immobilising trauma or surgery 6 weeks prior to the event, cancer diagnosis 3 months before or after the event, exposure to other sex hormones, congenital heart disease, drug overdose, occurrence of the event within 6 months of the woman's censoring date for the study. |
| Nwaru, 2021 ([40]) | Women aged 16-45 years who were registered with a general practice and had any asthma event (including diagnosis, hospitalisation, medication prescription) during the study period. Women were excluded in any year when they were pregnant. |
| Parker, 2016 ([23]) | Women who were registered with a general practice and had a singleton delivery during the study period. Women with pre-existing chronic hypertension requiring treatment with an antihypertensive were excluded. |

| First author, year (reference) | Study population description |
|---|---|
| | Cases: women with first-time pre-eclampsia during the study period.<br>Controls: women with no history of pre-eclampsia. |
| Pasvol, 2022 ([16]) | Women aged 15-49 years who were registered with a general practice during the study period. Women were censored at first recording of an event that would preclude contraception use (e.g. hysterectomy), first recording of HRT prescription, date they de-registered, or date of death. |
| Rees, 2016 ([24]) | Women aged 15-44 years who were registered with a general practice for which data could be linked to Hospital Episode Statistics.<br>Cases: women with a diagnosis of PCOS.<br>Controls: women with no diagnosis of PCOS, who remained at the same practice for at least the same duration from index date as their respective case. |
| Reddy, 2014 ([42]) | Women aged 12-55 years who were registered with a general practice during the study period (191 general practices). |
| Richardson, 2018 ([29]) | Women aged 18-55 years with a coded non-inflammatory potentially painful musculoskeletal condition who were registered with a general practice for which data could be linked to the Office for National Statistics and Hospital Episode Statistics (350 general practices).<br>Exclusion criteria: a cancer diagnosis at any time prior to the first day of opioid use or within the following 6 months and <1 year of records within the database prior to the first day of opioid prescription.<br>Cases: women starting a long-term opioid (at least three opioid prescriptions within 90 days from and including the first date of a new prescription) at the time of a musculoskeletal condition.<br>Controls: women starting a short-term opioid (maximum of two opioid prescriptions within 90-day period) at the time of a musculoskeletal condition. |
| Rowlands, 2000 ([17]) | Women aged 14-29 years permanently registered with a general practice that contributed data that met the quality standard set by the Office for National Statistics throughout the study period (357 general practices). |
| Seaman, 2003 ([34]) | Women aged 15-39 years who were registered with a general practice that contributed data deemed to be of research standard by the owners of the database (UK Medicines Control Agency). |
| Shawe, 2008 ([32]) | Women aged 15-44 years who were registered with a general practice that contributed data deemed to be of research standard (136 general practices).<br>Women were excluded if they had been sterilised or had a hysterectomy prior to 2001.<br>Cases: women with a code for type 1 or 2 diabetes prior to 2001 (excluded: women with a first code for diabetes after 2001 or with only a history of gestational diabetes).<br>Controls: women with no record of or treatment for diabetes at any time in their medical record. |
| Shorvon, 2022 ([35]) | Women aged 15-45 years who were registered with a general practice and who had received a prescription for an antiepileptic drug during the study period and a diagnosis of epilepsy or epileptic seizures in their medical record (294 general practices). |
| Smith, 2020 ([18]) | Women aged 15-49 years who gave birth to a single live infant between 2006-2015 and who were registered with a general practice (730 general practices). Women were excluded if they had multiple deliveries (e.g. twins), known miscarriage, termination or stillbirth, were registered with a practice which did not have acceptable computer use (ACU) or acceptable mortality rates (AMR) by the date of childbirth, or were registered at a practice for less than 6 months. |
| Smith, 2021 ([41]) | Women aged 15-23 years who participated in the Avon Longitudinal Study of Parents and Children (ALSPAC) and had linked primary care data at relevant time points (at least 18 months before and 6 months after their clinic visit or questionnaire completion), |

| First author, year (reference) | Study population description |
|---|---|
| Subramanian, 2022 ([19]) | Pregnant women aged 15-49 years who were registered with a general practice and had a record of delivery from linked Hospital Episode Statistics. Delivery records were excluded if they were duplicates, misclassified miscarriage, postnatal or antenatal record, or women were ineligible (not aged 15-49 or no minimum registration period of 1 year) or lost to follow-up within primary care at the delivery. |
| Syed, 2022 ([20]) | Women aged 16-55 years who were registered with a general practice who had a live birth recorded and data linked to Hospital Episode Statistics and the Office for National Statistics mortality register. Children had to be registered with the practice within 6 months after birth, with follow-up data collected until the child’s first birthday. |
| Wemakor, 2014 ([21]) | Women aged 15-45 years who were registered with a general practice with a high coverage of electronic prescribing data (≥70% of prescriptions could be recovered from the electronic system). Women had to be registered with the same general practice throughout the study period (246 general practices). |

PCOS, polycystic ovary syndrome.
